# Deep learning-based electrocardiographic differentiation of Takotsubo syndrome from acute anterior myocardial infarction: explainability, human-AI interaction, and a utilization bias hypothesis

**DOI:** 10.64898/2026.10.15.26365116

**Authors:** Eisuke Kagawa, Masaya Kato, Takayuki Hidaka, Eiji Kunita, Naoya Hironobe, Shogo Matsui, Yusuke Kobayashi, Yukimi Uotani, Tomoki Teramoto, Kanade Yoshii, Takanori Okumoto, Mikio Tani, Keigo Dote

**Affiliations:** Department of Cardiology, Hiroshima City North Medical Center Asa Citizens Hospital, Hiroshima, Japan; Director, Asa Medical Association Hospital, Hiroshima, Japan

**Keywords:** Takotsubo syndrome, acute myocardial infarction, electrocardiography, artificial intelligence, human-AI interaction, explainable artificial intelligence

## Abstract

**Background and Aims:** Artificial intelligence (AI) may help differentiate Takotsubo syndrome from acute anterior ST-segment elevation myocardial infarction (STEMI) on electrocardiograms (ECGs). We developed an ECG image-based model and evaluated whether AI assistance and visual explanations improved reader performance, overall and by reader background.

**Methods:** A self-supervised three-architecture deep learning ensemble was developed and externally validated using 12-lead ECG images from two institutions, recorded ≤14 days after onset regardless of reperfusion. A four-phase reader study compared unaided interpretation, a reference sheet, and AI assistance (Phases 1–3). In Phase 4, readers were randomized to the reference sheet alone, AI predictions, or predictions with gradient-weighted class activation mapping (Grad-CAM). Correctness was analyzed with generalized linear mixed-effects models.

**Results:** Cohorts comprised 832 development and 95 external validation participants. External validation accuracy was 0.914, and the area under the receiver operating characteristic curve was 0.970 (95% CI, 0.938–0.992), or 0.958 using first recorded ECGs. Among 70 readers, the estimated probability of correct diagnosis increased from 0.632 unaided to 0.864 with AI (odds ratio [OR], 3.71; P<0.001). In Phase 4, AI assistance improved accuracy vs reference sheet (OR, 4.28; 95% CI, 3.08–5.95), and Grad-CAM added value (OR, 1.53; 95% CI, 1.04–2.25). In exploratory analyses, less experienced readers gained more; with AI, non-cardiologists outperformed cardiologists. Mean AI-assisted reader accuracy remained below model accuracy.

**Conclusions:** An ECG image-based ensemble differentiated Takotsubo syndrome from anterior STEMI and improved reader performance, particularly with visual explanations. Experience-related differences emphasize the need for effective human-AI interaction.

**Structured Graphical Abstract:** Deep learning-based electrocardiographic differentiation of Takotsubo syndrome from acute anterior myocardial infarction: explainability, human-AI interaction, and a utilization bias hypothesis

**Graphical abstract legend:** ECG images from two hospitals supported development and external validation of a self-supervised, three-architecture ensemble. Seventy readers completed four phases. Phase 4 randomized readers to the reference sheet alone, with AI predictions, or with AI predictions plus Grad-CAM. Bars show mixed-model estimated probabilities of correct diagnosis (95% CI); the red dashed line indicates AI-alone accuracy on both 50-ECG sets. Exploratory subgroup odds ratios compare reader backgrounds in Phases 1 and 3.

**Abbreviations used in the graphic:** AI, artificial intelligence; AUC, area under the receiver operating characteristic curve; CI, confidence interval; ECG, electrocardiogram; Grad-CAM, gradient-weighted class activation mapping; MLP, multilayer perceptron; OR, odds ratio; STEMI, ST-segment elevation myocardial infarction; ViT, Vision Transformer.

**Alt text:** Three panels show ECG datasets from two hospitals, a three-model ensemble distinguishing Takotsubo syndrome from acute anterior STEMI (external validation AUC 0.970), and results for 70 readers. In randomized Phase 4, the estimated probability of correct diagnosis was 61% with a reference sheet alone, 84% with added AI predictions, and 89% with predictions plus heatmaps. In exploratory analyses, cardiologists’ unaided advantage reversed with AI assistance.

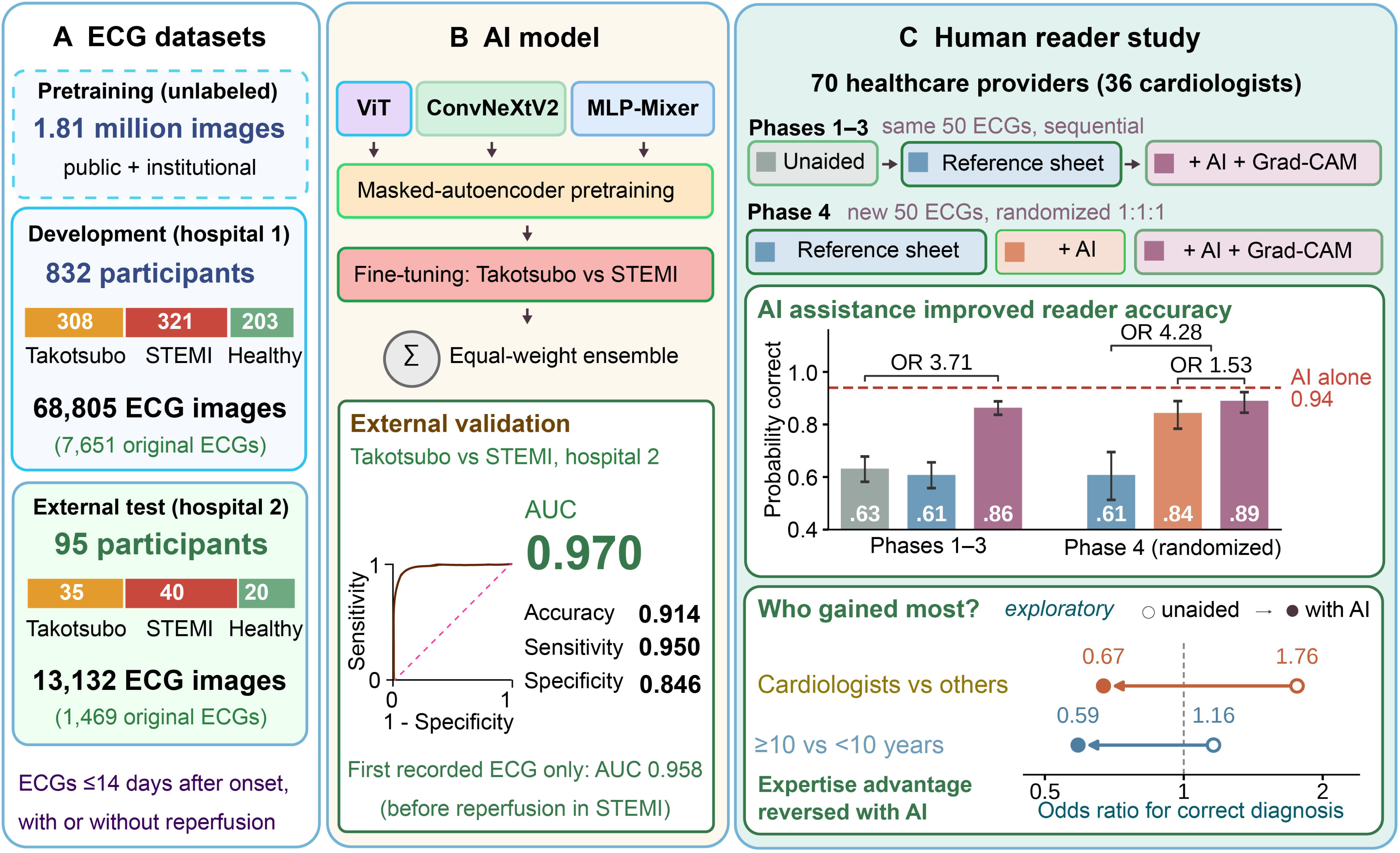

**Key Question:** Can a deep learning model applied to electrocardiogram images differentiate Takotsubo syndrome from acute anterior ST-segment elevation myocardial infarction, and does artificial intelligence (AI) assistance, with or without visual explanations, improve clinicians’ diagnostic accuracy?

**Key Finding:** In external validation, the ensemble achieved 0.914 accuracy and an area under the receiver operating characteristic curve of 0.970. Among 70 readers, AI assistance improved accuracy (randomized Phase 4 odds ratio 4.28), and heatmap explanations added benefit (odds ratio 1.53).

**Take-home Message:** An electrocardiogram image-based AI model differentiated Takotsubo syndrome from anterior myocardial infarction and improved readers’ accuracy, especially with visual explanations. Greater gains in less experienced readers suggest that its value may depend on effective human-AI interaction, not expertise alone.

## Introduction

Takotsubo syndrome, also known as stress cardiomyopathy and initially termed “Takotsubo cardiomyopathy” in Hiroshima, often presents with chest pain, electrocardiographic changes, and left ventricular wall-motion abnormalities that closely resemble acute myocardial infarction (AMI).^1–4^ In ST-segment elevation myocardial infarction (STEMI), prompt reperfusion therapy is essential for reducing mortality, whereas Takotsubo syndrome does not require similarly time-critical disease-specific treatment. Therefore, early differentiation between these conditions represents an important clinical challenge. Although invasive coronary angiography is often required to exclude obstructive coronary artery disease, patients with Takotsubo syndrome may present with conditions that increase procedural risk. Therefore, accurate identification of true Takotsubo syndrome may support consideration of noninvasive alternatives and help avoid unnecessary invasive coronary angiography in selected patients. Electrocardiography (ECG) is a rapid, noninvasive test routinely used in the initial evaluation of chest pain and is recommended within 10 minutes of first medical contact.^5^ Several studies have proposed ECG-based criteria to distinguish Takotsubo syndrome from AMI; however, their diagnostic performance remains suboptimal in real-world practice. This limitation is partly due to variability in clinical presentation, including spontaneous recanalization in AMI and temporal changes in ECG waveforms in both conditions, which can obscure distinguishing features.

Transthoracic echocardiography is also widely used and may aid in differentiation based on characteristic wall-motion patterns. However, accurate differentiation can remain challenging because of suboptimal imaging conditions in emergency settings. Consequently, Takotsubo syndrome is often diagnosed only after coronary angiography has been performed for suspected AMI.

Recent advances in artificial intelligence (AI), particularly deep learning, have enabled automated ECG analysis, with several studies demonstrating high diagnostic performance across various cardiovascular tasks.^6,7^ However, the clinical utility of such models depends not only on their predictive performance but also on their integration into clinical workflows and their potential to support human decision-making.^8^

The aims of this study were threefold: (1) to develop and externally validate an AI model for differentiating Takotsubo syndrome from acute anterior STEMI using ECGs obtained within 14 days after symptom onset, regardless of reperfusion status, and compare its performance with that of human readers; (2) to assess whether AI assistance improves reader diagnostic performance and whether visual explanations provide additional benefit; and (3) to explore whether performance gains and accuracy with AI assistance vary by cardiologist status and clinical experience, extending our previous work on an AI utilization gap.^8^

## Methods

### Study patients and data collection

This study included three subsets of participants: patients with acute anterior STEMI, patients with Takotsubo syndrome, and healthy volunteers. The three subsets were retrospectively identified at Hiroshima City North Medical Center Asa Citizens Hospital and Asa Medical Association Hospital. Patients diagnosed with acute anterior STEMI with a culprit lesion in the left-main trunk or left anterior descending artery (LAD), or with Takotsubo syndrome, between 1996 and 2024 were eligible for inclusion. The diagnosis of Takotsubo syndrome was based mainly on the Mayo criteria or the InterTAK Diagnostic Criteria, and patients with intracranial hemorrhage or brain trauma were also eligible for inclusion.^9,10^ Among eligible patients with Takotsubo syndrome, those who underwent coronary angiography or computed tomography were included in this study. Among eligible STEMI patients, 321 and 40 patients from Hiroshima City North Medical Center Asa Citizens Hospital and Asa Medical Association Hospital, respectively, were randomly sampled and included, based on a preliminary study indicating that these sample sizes would yield numbers of ECG images approximately balanced with those obtained if all candidate patients with Takotsubo syndrome were included. We reviewed the medical records of the study patients and collected ECGs. ECGs recorded within 14 days after symptom onset, with or without coronary revascularization, were labeled “Takotsubo” or “AMI,” whereas all ECGs recorded before symptom onset or at least 1 year after onset were labeled “non-takotsubo/AMI.” ECGs recorded between 14 days and 1 year after symptom onset were excluded. Healthy staff volunteers with ECGs recorded during routine health examinations and no relevant medical history were also included. For healthy volunteers, all recorded ECGs were collected and labeled “non-takotsubo/AMI.”

Twelve-lead ECG images were obtained by electronic export or photography of paper ECGs or screen displays; acquisition details are provided in the Supplementary Methods.

At Hiroshima City North Medical Center Asa Citizens Hospital, participants with Takotsubo syndrome, AMI, and healthy volunteers were assigned at the participant level to the training, validation, and pretraining-validation datasets in an approximate ratio of 75%, 20%, and 5%, respectively, such that each participant was included in only one dataset. An external test dataset was constructed from data collected at Asa Medical Association Hospital. Data from the former hospital were used for model development, whereas data from the latter hospital were used for external evaluation of the models and for the human diagnostic test. This study was approved by the Institutional Review Board of Hiroshima City North Medical Center Asa Citizens Hospital, which waived informed consent for patients, healthy volunteers, and readers, and was conducted in accordance with the Declaration of Helsinki.

### Pretraining datasets

For self-supervised pretraining, 1,754,276 ECG images were generated from the publicly available MIMIC-IV-ECG, A Large Scale 12-Lead Electrocardiogram Database for Arrhythmia Study, PTB-XL, and Georgia 12-Lead ECG Challenge Dataset.^11–14^ Our institutional dataset was also used for pretraining. Additional details are provided in the Supplementary Methods.

### Model development

We developed an ECG image-based deep learning ensemble model to distinguish Takotsubo syndrome from anterior STEMI with a culprit lesion in the left main trunk or LAD, with or without coronary reperfusion. Three backbone models—ViT, ConvNeXtV2, and MLP-Mixer—were pretrained using masked autoencoder-based self-supervised learning and subsequently fine-tuned, first for 3-class classification (Takotsubo syndrome, AMI, and non-Takotsubo/AMI) and then for binary classification (Takotsubo syndrome vs AMI). During inference, test-time augmentation (TTA) was applied where appropriate. The final ensemble prediction was generated by averaging logits from the three component models with equal weights.^15^ Details of the model architectures, pretraining, fine-tuning, checkpoint selection, and TTA are provided in the Supplementary Methods.

### Evaluation of models with external dataset

A predicted probability of ≥ 0.5 was used to classify an ECG as AMI. Performance metrics were assessed using accuracy, precision, sensitivity (recall), specificity, F1 score, expected calibration error (ECE), Brier score, and Brier skill score (BSS). Receiver operating characteristic (ROC) and precision-recall (PR) curves were constructed using the predicted probability of AMI, and the corresponding areas under the curves (AUCs) were calculated. The external dataset was used for evaluation only, without model recalibration or retraining. To approximate the initial diagnostic decision before coronary angiography, performance was also evaluated using images derived from each patient’s first recorded ECG (before reperfusion in AMI). Three same-architecture ensembles, each comprising three models, were compared with the final ensemble in an exploratory analysis (Supplementary Methods). Because multiple images were derived from each patient, 95% confidence intervals (CIs) were estimated using 5,000 patient-cluster bootstrap replicates stratified by diagnosis, and a sensitivity analysis weighted each image by the reciprocal of the patient’s number of images (Supplementary Methods).

### Human diagnostic test

In a four-phase reader study, volunteer healthcare providers with varying ECG experience interpreted ECGs without additional clinical information. Two 50-image ECG sets were randomly sampled from the test set: one for Phases 1 – 3 and another for Phase 4. Phases 1 – 3 involved unaided interpretation, a criteria-based reference sheet derived from prior reports^16–19^, and the reference sheet plus AI assistance, respectively (Supplementary Figure S1). AI assistance in Phase 3 included predictions from the three component models and their ensemble, and gradient-weighted class activation mapping (Grad-CAM) visualizations from ViT and ConvNeXtV2. In Phase 4, participants were assigned in a 1:1:1 ratio using stratified block randomization by cardiologist status and years of clinical experience to reference sheet alone, reference sheet plus AI predictions without Grad-CAM, or reference sheet plus AI predictions with Grad-CAM. For each ECG, readers provided a binary diagnosis (Takotsubo syndrome vs AMI). Accuracy, sensitivity, and specificity for AMI were calculated. Set composition and randomization details are provided in the Supplementary Methods.

### Statistical analysis

Continuous variables are presented as medians with first and third quartiles or as mean ± standard deviation, as appropriate. Categorical variables are presented as numbers and percentages. No missing data were observed in the baseline characteristics of the study patients. Diagnostic correctness at the ECG-response level was the primary outcome and was analyzed using binomial generalized linear mixed-effects models (GLMMs) with a logit link and random intercepts for participants and ECG cases. Fixed effects were phase for Phases 1 – 3 and randomized group for Phase 4. Prespecified contrasts compared Phase 2 vs Phase 1, Phase 3 vs Phase 2, and Phase 3 vs Phase 1. For all three pairwise comparisons among Phases 1 – 3, P values were adjusted using the Holm method, and 95% CIs for the odds ratios (ORs) were adjusted using the Bonferroni method. In Phases 1 – 3, separate exploratory GLMMs assessed interactions of phase with cardiologist status or clinical experience. In Phase 4, the primary comparison was reference sheet alone vs both AI-assisted groups combined; the secondary comparison was AI assistance with vs without Grad-CAM. Sensitivity analyses adjusted both the Phase 4 comparisons for cardiologist status and experience (< 10 vs ≥ 10 years). Accuracy was the proportion of correct diagnoses, with AMI as the positive class for sensitivity and specificity. Secondary participant-level analyses are described in the Supplementary Methods.

Sample size estimation for the human diagnostic study was based on participant-level accuracy in the pilot data. Because the comparison between AI assistance with and without Grad-CAM required the largest sample size, the target sample size was set at 20 participants per group. With 1:1:1 randomization in Phase 4, the planned total sample size was 60 participants. Additional details are provided in the Supplementary Methods.

We utilized Python 3.12 (Python Software Foundation, Delaware, USA) for model development, human test code, and statistical analyses. GLMMs were fitted using R software version 4.6 (R Foundation for Statistical Computing, Vienna, Austria) with the lme4 package.^20^ The TRIPOD+AI and STARD-AI reporting checklists are provided in the Supplementary Material.

## Results

### Baseline characteristics

Figure 1 summarizes the study. The development dataset comprised 832 participants (308 with Takotsubo syndrome, 321 with AMI, and 203 healthy controls) and 68,805 ECG images. The external validation dataset comprised 95 participants and 13,132 ECG images (Table 1). All patients with AMI underwent immediate coronary angiography, and primary percutaneous coronary intervention was attempted in 99%. Development-set partition sizes are shown in Supplementary Table S8.

**Figure 1.**
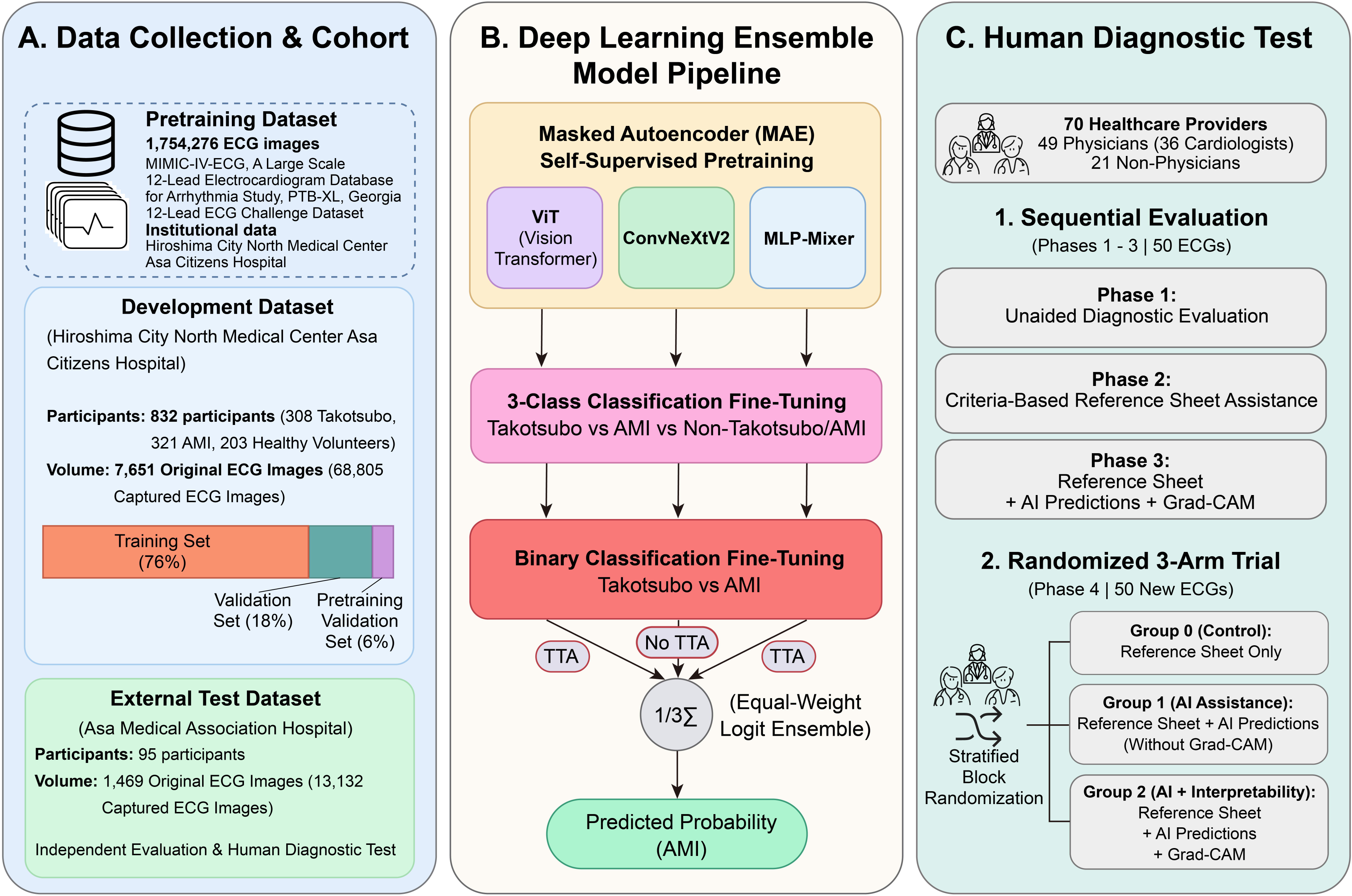
Study overview. This study comprised three stages: (A) Development and external-validation ECG image datasets were collected from two hospitals; (B) three self-supervised pretrained backbone models were fine-tuned and combined into the final ensemble model, which was evaluated on the external test dataset; and (C) the final model was tested against human readers in a 4-phase diagnostic study. (A) For self-supervised pretraining, 1,754,276 ECG images were generated from four publicly available databases (MIMIC-IV-ECG, A Large Scale 12-Lead Electrocardiogram Database for Arrhythmia Study, PTB-XL, and Georgia 12-Lead ECG Challenge Dataset). Additional institutional ECG images (Hiroshima City North Medical Center Asa Citizens Hospital) were used for pretraining and pretraining validation. Development and external-validation datasets were collected from two hospitals. Patients with acute anterior STEMI (left main or left anterior descending culprit) or Takotsubo syndrome, and healthy volunteers, contributed 12-lead ECGs; those recorded within 14 days after symptom onset were labeled AMI or Takotsubo, whereas those recorded before onset or at least 1 year after onset were labeled non-Takotsubo/AMI. The development dataset (832 participants) was divided at the participant level into training (n = 633; 76%), validation (n = 149; 18%), and pretraining-validation (n = 50; 6%) sets. (B) Three backbone models (Vision Transformer, ConvNeXtV2, and MLP-Mixer) were pretrained on unlabeled ECG images by MAE-based self-supervised learning, in which randomly masked image patches are reconstructed so that the models learn general ECG image representations without diagnostic labels. Each model was then fine-tuned for 3-class and subsequently binary classification, with TTA applied where appropriate, and the final prediction was obtained by equal-weight logit ensemble. Performance of the models was evaluated on the external test dataset. (C) Seventy healthcare providers (49 physicians, including 36 cardiologists, and 21 non-physicians) completed the 4-phase human diagnostic test. Phases 1 – 3 used a common 50-ECG set under unaided, reference sheet-assisted, and AI-assisted conditions, respectively. In Phase 4, participants interpreted a separate 50-ECG set after stratified block randomization in a 1:1:1 ratio to reference sheet alone (group 0), reference sheet + AI predictions without Grad-CAM (group 1), or reference sheet + AI predictions with Grad-CAM (group 2). AMI indicates acute myocardial infarction; STEMI, ST-segment elevation myocardial infarction; MAE, masked autoencoder; TTA, test-time augmentation; and Grad-CAM, gradient-weighted class activation mapping. Alt text: Study flow diagram showing ECG-image acquisition from two hospitals, participant-level development splits, self-supervised pretraining and fine-tuning of ViT, ConvNeXtV2, and MLP-Mixer, external validation of their ensemble, and a four-phase study involving 70 readers. Only Phase 4 used randomized support allocation.

**Table 1.** Baseline characteristics of the dataset.

|  | Development dataset |  |  | External validation dataset |  |  |
|---|---|---|---|---|---|---|
|  | Takotsubo syndrome | AMI | Healthy control | Takotsubo syndrome | AMI | Healthy control |
| N | 308 (37) | 321 (39) | 203 (24) | 35 (37) | 40 (42) | 20 (21) |
| Age, y | 77 [65 – 84] | 65 [54 – 75] | 47 [37 – 60] | 79 [70 – 86] | 74 [66 – 81] | 46 [39 – 54] |
| Male sex | 49 (16) | 238 (74) | 134 (66) | 11 (31) | 31 (78) | 12 (60) |
| Stress |  |  |  |  |  |  |
| Physical | 171 (55) |  |  | 29 (83) |  |  |
| Emotional | 128 (42) |  |  | 6 (17) |  |  |
| Happy heart syndrome | 9 (3) |  |  | 0 |  |  |
| Unknown | 9 (3) |  |  | 0 |  |  |
| Type |  |  |  |  |  |  |
| Atypical (non-apical type) | 14 (5) |  |  | 1 (3) |  |  |
| Infarct related artery |  |  |  |  |  |  |
| Left main trunk |  | 7 (2) |  |  | 1 (2) |  |
| LAD |  | 314 (98) |  |  | 39 (98) |  |
| Original ECG, n | 1729 | 3531 | 2391 | 345 | 651 | 473 |
| Captured ECG, n | 15559 | 31731 | 21515 | 3104 | 5779 | 4249 |

Data were presented as n (%) or median (interquartile range).
AMI indicates acute myocardial infarction; LAD, left anterior descending artery; and ECG, electrocardiogram.

### External validation performance of the AI models

Representative ECG images with corresponding model predictions and visualization for Takotsubo syndrome and AMI are shown in Supplementary Figure S2. The final ensemble achieved an accuracy of 0.914, sensitivity of 0.950, specificity of 0.846, and ROC AUC of 0.970; additional performance metrics are shown in Table 2, Figure 1, and Figure 2.

**Figure 2.**
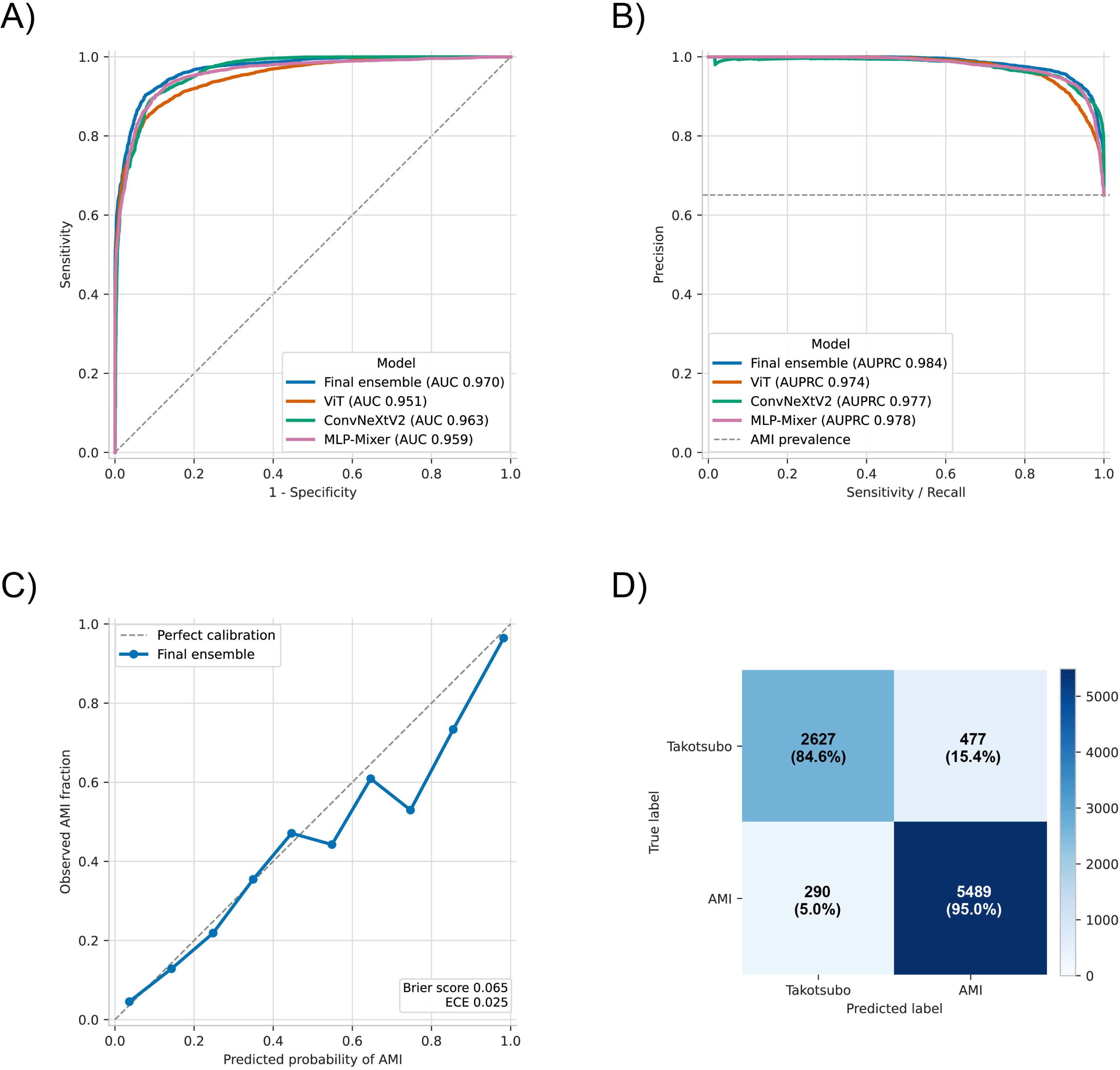
Performance of the final ensemble model. ROC curves (A) and PR curves (B) of the final ensemble model and its three component models for predicting AMI. The calibration plot (C) shows the agreement between predicted probability and observed proportion of AMI for the final ensemble model. The confusion matrix (D) shows image-based classification by the final ensemble model at a predicted AMI probability threshold of 0.5. Each cell shows the number of ECG images and the corresponding row percentage, calculated using the total number of images in each true diagnostic class as the denominator. AMI indicates acute myocardial infarction; ECG, electrocardiogram; ROC, receiver operating characteristic; and PR, precision-recall. Alt text: Four panels show ROC and precision-recall curves for the ensemble and its component models, an ensemble calibration plot, and a confusion matrix. The ensemble achieved external validation ROC AUC 0.970 and accuracy 0.914. Confusion-matrix percentages use each true diagnostic class as denominator.

**Table 2.** External validation performance of the AI models.

|  | Accuracy | Precision | Recall (sensitivity) | Specificity | F1 score | ROC AUC | PR AUC | ECE | Brier score | BSS |
|---|---|---|---|---|---|---|---|---|---|---|
| Component model (backbone) |  |  |  |  |  |  |  |  |  |  |
| ViT | 0.882 | 0.908 | 0.912 | 0.828 | 0.910 | 0.951 | 0.974 | 0.094 | 0.094 | 0.584 |
| ConvNeXtV2 | 0.899 | 0.892 | 0.962 | 0.782 | 0.925 | 0.963 | 0.977 | 0.039 | 0.076 | 0.666 |
| MLP-Mixer | 0.907 | 0.919 | 0.939 | 0.847 | 0.929 | 0.959 | 0.978 | 0.069 | 0.081 | 0.642 |
| Final ensemble model | 0.914 | 0.920 | 0.950 | 0.846 | 0.935 | 0.970 | 0.984 | 0.025 | 0.065 | 0.712 |
| First recorded ECG only (836 images) |  |  |  |  |  |  |  |  |  |  |
| Final ensemble model | 0.864 | 0.822 | 0.951 | 0.763 | 0.882 | 0.958 | 0.965 | 0.093 | 0.097 | 0.609 |

Values are image-level estimates using all 8,883 Takotsubo and AMI images in the external validation dataset, except for the first recorded ECG analysis, which used 836 images derived from each patient’s first recorded ECG (before reperfusion in AMI); component-model results for this analysis are shown in Supplementary Table S3.
AI indicates artificial intelligence; ROC, receiver operating characteristic; AUC, area under the curve; PR, precision-recall; ECE, expected calibration error; and BSS, Brier skill score.

External ablation analyses showed improved performance with ImageNet initialization and the highest performance with self-supervised pretraining, compared with training from scratch. TTA further improved ViT and MLP-Mixer performance; it was not used for ConvNeXtV2 (Supplementary Table S1). The final ensemble had the highest observed ROC AUC (0.970 vs 0.952, 0.965, and 0.959 for the ViT-only, ConvNeXtV2-only, and MLP-Mixer only ensembles, respectively; Supplementary Table S2).

Using images derived from each patient’s first recorded ECG in the external validation dataset – before reperfusion in AMI and after the index event in Takotsubo syndrome – the ensemble achieved an accuracy of 0.864, sensitivity of 0.951, specificity of 0.763, and ROC AUC of 0.958 (Table 2; Supplementary Table S3 and Figure S3). For the full external test dataset, patient-cluster bootstrap 95% CIs were 0.866 – 0.954 for accuracy and 0.938 – 0.992 for ROC AUC, and weighting each patient equally yielded an accuracy of 0.903 and ROC AUC of 0.968 (Supplementary Table S4).

### Human test

Seventy participants completed the reader study, including 36 cardiologists (51%); participants’ characteristics were balanced across randomized groups (Supplementary Table S5). The ensemble achieved an accuracy of 0.940 on each of the two 50-ECG sets (Figure 3 and Table 3). ECG-set composition is shown in Supplementary Table S6.

**Figure 3.**
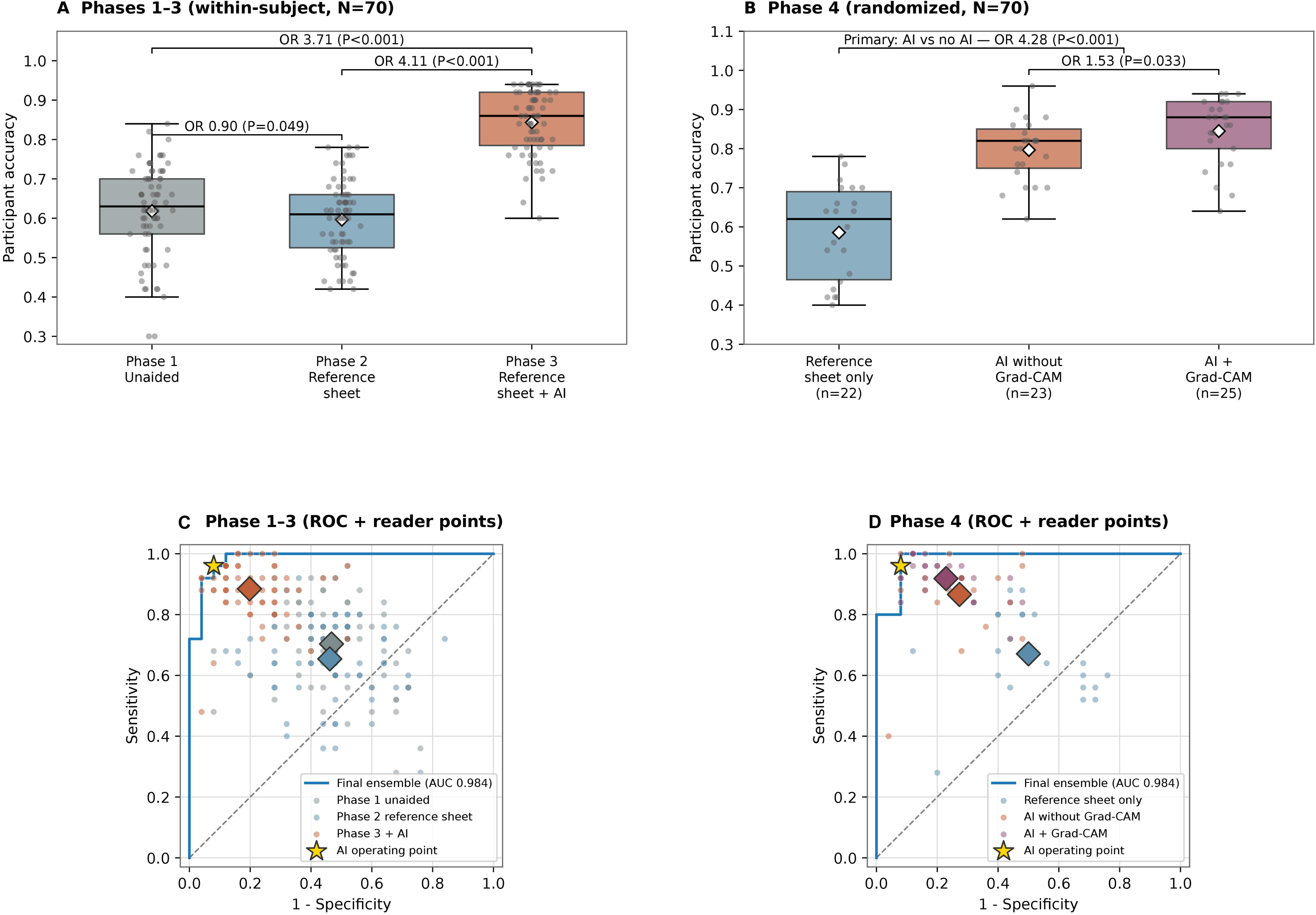
Human reader performance with and without AI assistance. Seventy participants interpreted 12-lead ECGs to differentiate Takotsubo syndrome from acute anterior STEMI. (A) Sequential interpretation of the same 50 ECGs without assistance (Phase 1), with a reference sheet (Phase 2), and with the reference sheet plus AI predictions and Grad-CAM visualization (Phase 3). (B) Phase 4 performance on a different set of 50 ECGs. Readers were randomized to the reference sheet alone (n = 22), the reference sheet plus AI predictions without Grad-CAM (n = 23), or the reference sheet plus AI predictions with Grad-CAM (n = 25). In A and B, boxes indicate the interquartile range, horizontal lines indicate medians, whiskers extend to the most extreme observations within 1.5 times the interquartile range of the box edges, white diamonds indicate means, and small grey dots represent individual readers. ORs and P values in A and B were obtained from generalized linear mixed models with random intercepts for readers and ECG cases. P values in panel A were Holm-adjusted for the three pairwise phase comparisons. (C and D) AI ROC curves and reader operating points for the Phases 1 – 3 test set (C) and the Phase 4 test set (D), with AMI as the positive class. Blue curves represent the final ensemble model; small coloured dots represent individual readers; large coloured diamonds indicate group mean operating points; and yellow stars indicate the AI operating point at an AMI probability threshold of 0.5. AI indicates artificial intelligence; AMI, acute myocardial infarction; AUC, area under the curve; CI, confidence interval; ECG, electrocardiogram; Grad-CAM, gradient-weighted class activation mapping; OR, odds ratio; ROC, receiver operating characteristic; and STEMI, ST-segment elevation myocardial infarction. Alt text: Four panels compare reader accuracy across sequential Phases 1–3 and randomized support groups in Phase 4, and show AI ROC curves with reader operating points for the two ECG sets. AI support improved accuracy; Grad-CAM provided an additional benefit in Phase 4.

**Table 3.** Human diagnostic performance.

|  | Comparison | N | Mean accuracy | Estimated probability (95% CI) | Odds ratio (95% CI) | P |
|---|---|---|---|---|---|---|
| Phase 1 (unaided) | Reference | 70 | 0.618 | 0.632 (0.582 – 0.678) |  |  |
| Phase 2 (reference sheet) | Phase 1 | 70 | 0.596 | 0.608 (0.558 – 0.656) | 0.90 (0.80 – 1.02) | 0.049 |
| Phase 3 (reference sheet + AI) | Phase 1 | 70 | 0.843 | 0.864 (0.837 – 0.888) | 3.71 (3.21 – 4.29) | < 0.001 |
|  | Phase 2 |  |  |  | 4.11 (3.55 – 4.75) | < 0.001 |
| Phase 4 (randomized) |  | 70 |  |  |  |  |
| Group 0 (reference sheet) | Reference | 22 | 0.585 | 0.608 (0.513 – 0.695) |  |  |
| Group 1 (AI without Grad-CAM) |  | 23 | 0.797 | 0.844 (0.784 – 0.889) |  |  |
| Group 2 (AI with Grad-CAM) |  | 25 | 0.845 | 0.890 (0.845 – 0.923) |  |  |
| AI assistance vs reference sheet | Primary |  |  |  | 4.28 (3.08 – 5.95) | < 0.001 |
| AI + Grad-CAM vs AI without Grad-CAM | Secondary |  |  |  | 1.53 (1.04 – 2.25) | 0.033 |
| The final AI model |  |  |  |  |  |  |
| Phase 1 – 3 set |  |  | 0.940 |  |  |  |
| Phase 4 set |  |  | 0.940 |  |  |  |

Estimated probability = subject-specific GLMM estimates; participant-level accuracy = raw per-reader mean; odds ratio, 95% CI from GLMM. For all three pairwise comparisons among Phases 1 – 3, P values were adjusted using the Holm method, and 95% CIs for the ORs were adjusted using the Bonferroni method.
AMI indicates acute myocardial infarction; GLMM, generalized linear mixed model; and CI, confidence interval.

In Phases 1 – 3, the GLMM-estimated probability of correct diagnosis was 0.632 (95% CI, 0.582 – 0.678) in Phase 1 (unaided), 0.608 (0.558 – 0.656) in Phase 2 (reference sheet), and 0.864 (0.837 – 0.888) in Phase 3 (reference sheet + AI). The accuracy in Phase 3 was significantly higher than in Phase 1 (odds ratio (OR) 3.71; 95% CI, 3.21 – 4.29, P < 0.001) and in Phase 2 (OR 4.11, 3.55 – 4.75, P < 0.001), whereas the reference sheet alone slightly reduced accuracy relative to unaided interpretation (Phase 2 vs Phase 1, OR 0.90, 0.80 – 1.02, P = 0.049). The participant-level analysis was concordant (Friedman P < 0.001, Phase 3 vs Phase 1 and Phase 3 vs Phase 2, both P < 0.001) (Figure 3). At the reader level, accuracy increased from Phase 1 to Phase 3 in 68 of 70 readers and from Phase 2 to Phase 3 in 69 readers, whereas it decreased from Phase 1 to Phase 2 in 37 readers (Figure 4).

**Figure 4.**
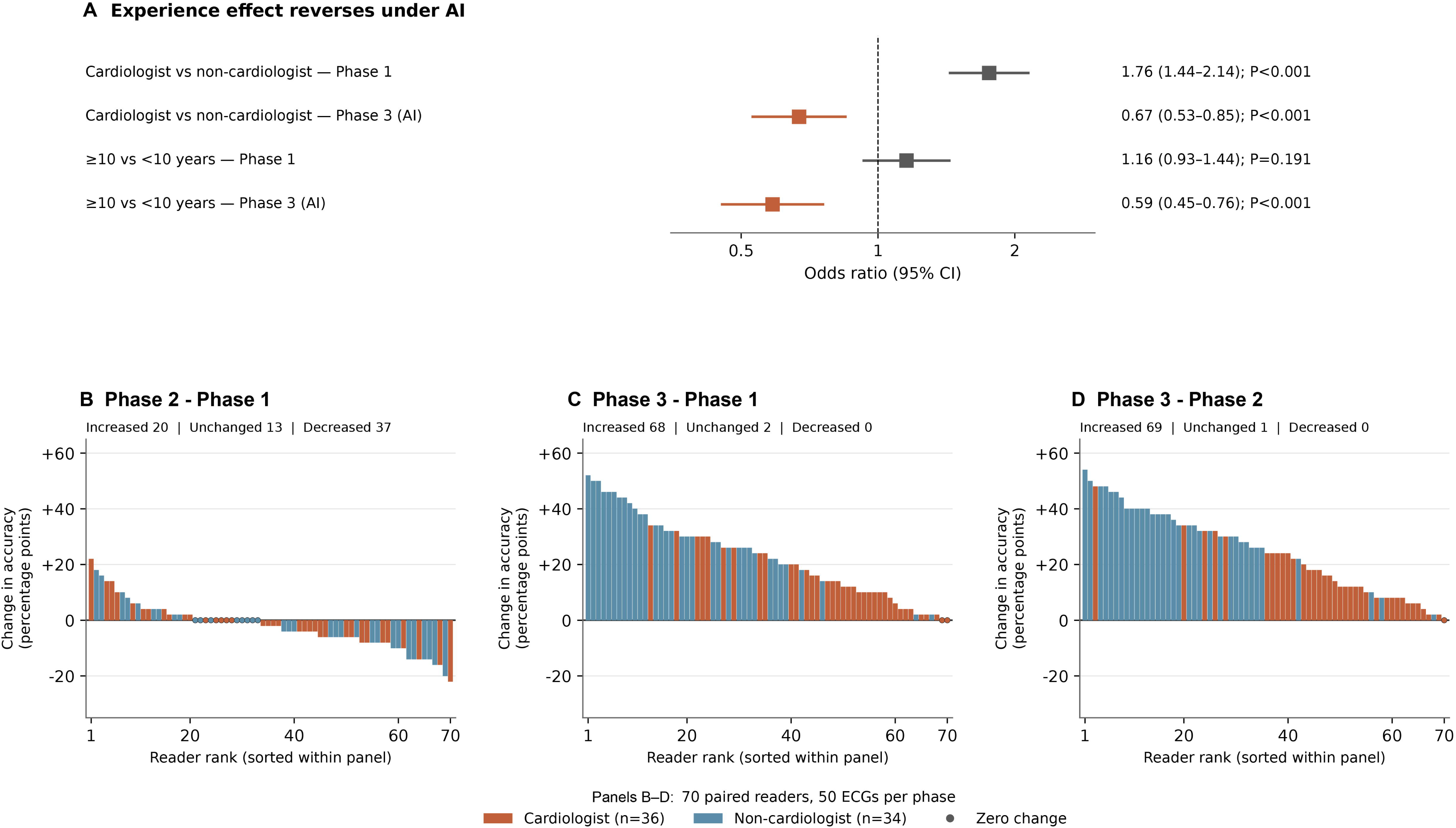
Reader background, clinical experience, and reader-level changes in accuracy. (A) Exploratory within-phase association of cardiologist status and clinical experience with diagnostic correctness in Phases 1 and 3. Squares indicate ORs and horizontal lines indicate 95% CIs; the dashed line indicates an OR of 1. ORs, 95% CIs, and P values were obtained from generalized linear mixed models with random intercepts for readers and ECG cases. (B – D) Reader-level changes in observed accuracy from Phase 1 to Phase 2 (B), Phase 1 to Phase 3 (C), and Phase 2 to Phase 3 (D). Each bar represents one reader’s paired change in accuracy, expressed in percentage points; positive values indicate higher accuracy in the later phase. Orange and blue indicate cardiologists and non-cardiologists, respectively. Dots on the zero line indicate unchanged accuracy. Readers are sorted independently in descending order within each panel; the same horizontal position therefore does not necessarily represent the same reader across panels. AI indicates artificial intelligence; CI, confidence interval; ECG, electrocardiogram; and OR, odds ratio. Alt text: A forest plot shows exploratory association of cardiologist status and experience with correctness in Phases 1 and 3. Three waterfall plots show each reader’s change in accuracy for Phase 2 minus Phase 1, Phase 3 minus Phase 1, and Phase 3 minus Phase 2, with cardiologist status indicated by colour.

In the randomized Phase 4, 22, 23, and 25 participants were assigned to the reference sheet-only, AI-without-Grad-CAM, and AI-with-Grad-CAM groups, respectively. The model-estimated probability of correct diagnosis was 0.608 (0.513 – 0.695), 0.844 (0.784 – 0.889), and 0.890 (0.845 – 0.923), respectively. In the prespecified primary comparison, participants with AI assistance had significantly higher accuracy than those with the reference sheet alone (OR 4.28, 95% CI 3.08 – 5.95, P < 0.001). In the prespecified secondary comparison, AI assistance with Grad-CAM was superior to AI assistance without Grad-CAM (OR 1.53, 1.04 – 2.25, P = 0.033). Additional pairwise comparisons among the three Phase 4 groups are shown in Supplementary Figure S4. Participant-level analyses were concordant (Kruskal-Wallis P < 0.001; AI vs no AI, P < 0.001; Grad-CAM vs no-Grad-CAM, P = 0.027) (Figure 3B). Stratification-adjusted estimates were similar (Supplementary Table S7).

In exploratory analyses of Phases 1 – 3, the benefit of AI assistance differed by participants’ experience. Interaction terms were significant for phase x experience group, phase x years of experience, and phase x cardiologist status (all P < 0.001). In Phase 1, cardiologists were more accurate than non-cardiologists (OR 1.76, 95% CI 1.44 – 2.14, P < 0.001). With AI assistance (Phase 3), less experienced participants achieved higher accuracy than more experienced participants (≥ 10 vs < 10 years: OR 0.59, 95% CI 0.45 – 0.76, P < 0.001), and non-cardiologists outperformed cardiologists (cardiologists vs non-cardiologists: OR 0.67, 95% CI 0.53 – 0.85, P < 0.001). Thus, although all groups improved with AI assistance, the incremental benefit was greatest among less experienced participants (Figure 4A; Supplementary Figures S5 and S6). Reader-level changes in observed accuracy are shown in Figure 4B–D.

## Discussion

In this study, we developed an ECG-based deep learning model to differentiate Takotsubo syndrome from acute anterior STEMI, including patients with or without coronary recanalization. Through self-supervised pretraining and integration of transformer-based, convolutional neural network–based, and multilayer perceptron–based models, the ensemble achieved robust diagnostic performance on an external validation dataset and exceeded mean reader accuracy on the reader-study ECG sets. In the reader study, AI assistance improved diagnostic accuracy, and adding Grad-CAM visualization provided further benefit. Exploratory analyses suggested that both improvement and AI-assisted accuracy varied by cardiologist status and clinical experience. These findings support prospective evaluation of AI-assisted ECG interpretation as an adjunct to clinical assessment. Its clinical impact, however, may depend on how effectively it is incorporated into human decision-making.

Several prior studies have addressed the differentiation of Takotsubo syndrome from acute myocardial infarction using ECG-based criteria or machine learning approaches.^16–19,21,22^ Shimizu et al. showed that machine learning using automatically extracted ECG parameters could distinguish Takotsubo syndrome from anterior myocardial infarction, and Hakansson et al. subsequently demonstrated in a larger registry-based cohort that an ECG-based machine learning model outperformed cardiologists using conventional ECG criteria. However, these studies focused mainly on model performance and did not fully address input usability, temporal ECG changes after symptom onset, or the effect of AI assistance on human diagnostic performance. Models requiring structured signal data or device-specific automated measurements may be difficult to apply across manufacturers or to ECGs archived or shared as images. By treating ECGs as images, our approach may be applicable to real-world formats, including photographs of paper-or display-based ECGs. The use of ECGs obtained within two weeks of symptom onset in both Takotsubo syndrome and anterior AMI, including post-reperfusion AMI patients, captured temporal changes encountered in routine clinical practice and allowed inclusion of patients with delayed presentation. Our human diagnostic test showed that AI assistance improved accuracy beyond unaided interpretation and conventional criteria-based support, while also revealing a potential utilization gap according to clinical experience.

Without TTA, self-supervised pretraining improved individual-model ROC AUCs by 0.17 – 0.23 relative to training from scratch. The final ensemble improved AUC by 0.007 – 0.019 over its components. It also had the highest observed AUC among the four ensemble configurations evaluated (Supplementary Table S2), supporting a potential benefit of architectural diversity for this task. Clinicians interpret ECGs by integrating local waveform morphology, inter-lead relationships, and overall spatial patterns. ConvNeXtV2’s convolutional inductive bias favors local morphological feature extraction, whereas ViT’s global self-attention supports modeling of long-range spatial relationships, potentially including relationships across ECG leads.^23,24^ MLP-Mixer integrates information across image patches and feature channels.^25^ These differences in inductive biases and information processing may provide complementary representations of ECG patterns, although the features underlying the ensemble gain were not directly assessed. TTA further improved discrimination for ViT and MLP-Mixer (Supplementary Table S1). These findings are consistent with prior work supporting ensembles for complex classification tasks.^26,27^

Overlapping ECG findings and temporal changes, including after spontaneous recanalization or coronary revascularization, complicate differentiation. Echocardiography may also be limited by suboptimal image acquisition, restricted patient positioning, and anatomical variations such as a wrapped LAD. These challenges support evaluating ECG models across the first two weeks after symptom onset, including after reperfusion. Performance was largely maintained when only each patient’s first recorded ECG was analyzed (ROC AUC 0.958), which corresponds to the initial diagnostic decision before coronary angiography.

One possible explanation for the superior performance of the AI model relative to human readers supported by criteria derived from prior reports is that AI models can capture subtle and complex ECG image patterns that are difficult to formalize as explicit diagnostic criteria. Traditional approaches rely on predefined features, such as ST-segment elevation/depression and T-wave inversion, as well as heuristic rules, which may not fully account for the variability observed in real-world ECGs. In contrast, deep neural network models can learn high-dimensional representations directly from ECG image data, enabling them to detect patterns that may not be readily apparent to human observers.

However, high predictive performance alone does not guarantee clinical impact; AI models must also be adopted and effectively used in real-world practice. Although many prior studies have demonstrated impressive model performance under experimental conditions, the integration of AI into clinical workflows and its influence on human decision-making remain insufficiently explored. Our human diagnostic test, which included 70 participants, represents one of the larger ECG reader studies evaluating AI-assisted interpretation and provides clinically relevant insights into AI-human interaction across a broad range of clinical experience. AI assistance significantly improved diagnostic accuracy compared with both unaided interpretation and conventional criteria-based support using a reference sheet derived from prior reports. Notably, randomized Phase 4 directly evaluated the incremental value of visual explanations: adding Grad-CAM heatmaps improved reader accuracy compared with AI assistance without these visualizations. This finding represents one of the relatively few examples in which an explainability component was associated with a measurable improvement in human diagnostic performance. These visual explanations may have encouraged participants to more readily incorporate AI predictions into their decisions, enhanced their own diagnostic interpretation, or both. However, even with AI assistance, mean reader accuracy remained below that of the AI model alone (0.94); only one reader exceeded the model’s accuracy, achieving 0.96 in Phase 4. This finding underscores that the existence of a high-performance model does not necessarily ensure its full utilization by human users. Further studies are needed to determine how the usability, interpretability, and implementation of AI-based decision-support tools, including their user interface and user experience, can be optimized to facilitate the appropriate use of AI predictions in clinical decision-making.

A similar pattern of smaller gains and lower AI-assisted accuracy among more experienced readers was observed in our previous study of AI-assisted prediction of elevated B-type natriuretic peptide levels from chest radiographs.^8^ Taken together with the superior performance of the model itself, this finding may be regarded as one form of progress in medical AI, yet it also carries a certain bittersweet irony. In this narrowly defined task, a data-driven model outperformed familiar human-centered approaches grounded in prior ECG studies and accumulated clinical experience. This echoes Sutton’s “Bitter Lesson”: throughout the history of AI research, major breakthroughs have often emerged not from encoding ever more human knowledge into systems, but from general methods capable of exploiting increasing computational resources.^28^

Across our two reader studies, this paradoxical pattern was characterized by three features: (1) AI assistance improved accuracy in both more and less experienced groups, with larger gains among less experienced readers; (2) the unaided advantage associated with cardiologist status in the present study and greater clinical experience in our previous study reversed with AI assistance; and (3) mean AI-assisted reader accuracy nevertheless remained below that of the AI model alone.^8^ Taken together, these findings suggest that conventional clinical expertise does not necessarily translate into effective use of AI assistance and that the relative performance of reader groups can reverse under AI support. These observations motivate a utilization bias hypothesis: greater reliance on judgements grounded in prior expertise may limit the effective incorporation of AI predictions, contributing to smaller performance gains and lower AI-assisted accuracy. This paradoxical pattern among more experienced readers may reflect a tendency to place greater weight on their own judgements than on AI predictions, potentially consistent with anchoring on prior judgements or cognitive entrenchment associated with expertise. These mechanisms were not directly assessed. The paradoxical pattern observed in our study may arise not only in AI-assisted decision-making but also in other fields, reflecting a possible tension between established expertise and adaptation to new tools or approaches. Although individual AI models may be rapidly superseded by more capable alternatives, challenges in their effective use, including the proposed utilization bias, may persist across successive generations of tools. At the same time, overreliance on AI assistance may impair unaided performance when users are subsequently required to complete similar tasks without AI support.^29^ Although AI tools can improve performance on narrowly defined tasks that are amenable to a divide-and-conquer approach, overreliance on them may contribute to the erosion of human skills. Traditional learning and human-centered approaches may help mitigate this risk. Even if more powerful AI agents or artificial general intelligence-like systems eventually emerge as potential “silver bullets,” a reasonable equilibrium may lie in combining effective use of such tools with continued human-centered training that encourages openness to new approaches, while pursuing sustained, incremental improvements in how the two are integrated.

### Study limitations

Several limitations should be considered. This was a retrospective study, and although an external validation dataset was included, further prospective validation is required to assess generalizability. The datasets were derived from specific institutions, and differences in patient populations, ECG acquisition methods, and clinical practice patterns may limit broader applicability. Performance was evaluated at the image level with multiple images per patient; however, patient-cluster bootstrap CIs and equal weighting of patients yielded similar estimates. The analysis was based on ECG data alone and did not incorporate clinical information, laboratory data, or imaging findings. Incorporating such information may further improve diagnostic performance. The human diagnostic test was conducted under controlled conditions and may not fully reflect real-world clinical workflows. Repeated ECG exposure in Phases 1 – 3 and patients shared between the two sets may have introduced learning or recall effects. Phase 4 randomization was expected to balance residual learning effects across groups. Finally, participants’ trust in AI and their reasons for accepting or rejecting AI predictions were not directly assessed.

## Conclusions

We developed an ECG image-based ensemble deep learning model that accurately differentiated Takotsubo syndrome from acute anterior STEMI. AI assistance improved reader accuracy, with additional benefit from visual explanations. Exploratory analyses suggested that both performance gains and AI-assisted accuracy varied by reader background and clinical experience. AI-supported ECG interpretation may enhance clinical decision-making in challenging diagnostic scenarios; further studies should evaluate performance in real-world clinical practice and address potential utilization gaps.

## Data Availability

The institutional electrocardiograms are availabe upon reasonable request to the authors. Other source codes, models, and sample codes are available online at GitHub.

https://github.com/ekagawa007/tako_ami_pytorch

## Acknowledgements

We sincerely thank all participants for their contribution to this study. During model development and manuscript preparation, the authors used ChatGPT (OpenAI, San Francisco, CA) and Claude (Anthropic, San Francisco, CA) to assist with Python coding and English-language editing. All AI-assisted code was reviewed and verified by the authors. Eisuke Kagawa had full access to all study data and takes responsibility for the integrity of the data and the accuracy of the analysis. The authors reviewed and approved the final text and take full responsibility for the content of the manuscript.

## Funding

There is no financial support for this study.

## Disclosure of Interest

None declared.

## Data availability statement

The institutional ECG data underlying this article cannot be shared publicly to protect patient privacy and will be shared on reasonable request to the corresponding author, subject to institutional approval. The public ECG databases used for pretraining are available from PhysioNet.^11–14^ The training code, trained models, and example inference code are available on GitHub (https://github.com/ekagawa007/takotsubo_ami_ai_ecg). Software version 0.4.0 is archived at https://doi.org/10.5281/zenodo.23763944. Trained model weights version 0.2.0 are archived at https://doi.org/10.5281/zenodo.23666262 and are also distributed at https://huggingface.co/ekagawa/takotsubo-ami-ai-ecg.

## Consent

The Institutional Review Board of Hiroshima City North Medical Center Asa Citizens Hospital approved this study and waived the requirement for informed consent from patients, healthy volunteers, and readers (08-1-42).

## Supplementary Material

Supplementary Methods

Supplementary Tables S1 – S8

Supplementary Figures S1 – S6

TRIPOD+AI checklist

STARD-AI checklist

References 30 - 35

## Notes

### Competing Interest Statement

The authors have declared no competing interest.

### Author Declarations

Ethical committee of North Medical Center Hiroshima Citizens Hospital and Asa Medical Association Hospital gave ethical approval for this work.

### Summary of Updates

Due to the change in the target journal, we have updated the figures and tables and added a graphical abstract to the manuscript.

