## Supplementary Materials for "Deep learning-based electrocardiographic differentiation of Takotsubo syndrome from acute anterior myocardial infarction: explainability, human-AI interaction, and a utilization bias hypothesis"

**Table of Contents:**

Supplementary Methods

Supplementary Tables S1 – S8

Supplementary Figures S1 – S6

Supplementary TRIPOD+AI checklist

Supplementary STARD-AI checklist

### Supplementary Methods

#### ECG collection

ECGs were recorded using electrocardiographs from multiple manufacturers, including Nihon Kohden (Tokyo, Japan), Fukuda Denshi (Tokyo, Japan), GE Healthcare (Chicago, IL, United States), and Philips (Amsterdam, the Netherlands). ECGs were collected as image data. ECGs were obtained in any format containing all 12 leads, including 6-channel/2-column, 3-channel/4-column, and 12-channel/1-column layouts. ECGs stored electronically in the electronic medical record system were obtained either by image export or by direct photography of the display. ECGs recorded on paper before implementation of the electronic medical record system were obtained by direct photography of the recording paper. In addition to the original image capture or frontal photograph, all ECGs were also photographed from eight additional viewing angles: up, down, left, right, and the four diagonal directions. Photographs were obtained using three Sony cameras: NEX-7, DSC-RX100M3, and ILCE-7RM2 (Sony, Tokyo, Japan).

In Japan, employers are required to provide annual health checkups for full-time employees and for part-time or temporary workers whose weekly working hours are at least three quarters of those of full-time staff, and electrocardiography is often performed as part of these examinations. Healthy staff volunteers who had one or more ECGs recorded at the two hospitals and had no specific medical history, including myocardial infarction or Takotsubo syndrome, were enrolled in this study.

**Quality control and diagnostic verification**

All institutional ECG images, clinical diagnoses, and image labels were checked by at least two investigators: Eisuke Kagawa and one or two of the following: Shogo Matsui, Yusuke Kobayashi, Tomoki Teramoto, Kanade Yoshii, Takanori Okumoto, or Mikio Tani. Images of insufficient quality were discarded. Patients diagnosed with Takotsubo syndrome or acute anterior STEMI were reviewed against the eligibility and diagnostic criteria specified in the main Methods; those who did not meet these criteria were excluded.

**Software environment**

At the completion of model training, the recorded software environment included PyTorch 2.7.1+cu128, torchvision 0.22.1+cu128, Hugging Face Transformers 4.57.1, and timm 1.0.21.

**Pretraining datasets**

Self-supervised pretraining datasets were constructed as follows. A total of 1,754,276 ECG images (480 × 480 pixels) in 6-channel/2-column and 3-channel/4-column formats were generated from the publicly available MIMIC-IV-ECG, A Large Scale 12-Lead Electrocardiogram Database for Arrhythmia Study, PTB-XL, and Georgia 12-Lead ECG Challenge Dataset.[11-14] Of these, 23,974 images (1.4%) were used as the validation set for pretraining. In addition, the training and pretraining-validation datasets of our original dataset were used for pretraining and pretraining validation, respectively. In total, approximately 1.81 million ECG images were used for self-supervised pretraining.

**Self-supervised pretraining of ViT (MAE)**

Hyperparameters were selected by grid search. We performed self-supervised pretraining (SSL) using a Masked Autoencoder Vision Transformer (ViT-MAE; facebook/vit-mae-base) implemented with the Hugging Face Transformers library (ViTMAEForPreTraining and AutoImageProcessor). [24,30-32] All ECG images were treated as RGB images and processed at an input size of 480 x 480 pixels using the pretrained image processor, including resizing and normalization followed by tensor conversion. During pretraining, we applied mild augmentation consisting of random resized cropping to 480 x 480 pixels with a scale range of 0.8-1.0; no flips were used because they are inappropriate for ECG morphology.

Pretraining used two image sources: (1) an external ECG image dataset for pretraining and (2) our original ECG image dataset. Training and validation datasets were created by concatenating the respective source-specific datasets.

Model optimization was conducted using the Hugging Face Trainer framework with custom dataloader hooks.[31] Training was run for up to 100 epochs with a batch size of 128 (evaluation batch size 16). We used a cosine learning-rate schedule with warmup (warmup ratio 0.05) and enabled bfloat16 mixed precision. Model checkpoints were saved at each epoch. To select pretrained weights for downstream supervised fine-tuning, we evaluated reconstruction loss on a held-out validation set for each saved checkpoint. No data augmentation was applied for validation images. Each checkpoint was loaded, and the mean validation reconstruction loss across batches was recorded. Train and validation loss curves were evaluated, and the checkpoint with the lowest validation reconstruction loss and no evidence of overfitting based on the training/validation reconstruction loss curves was selected for the pretrained initialization for downstream fine-tuning.

**Self-supervised pretraining of ConvNeXtV2 (FC-MAE)**

We performed self-supervised pretraining of a ConvNeXtV2-Base backbone using a fully convolutional masked autoencoding framework (FC-MAE) implemented in PyTorch using timm.[23] The pretraining objective was masked image reconstruction with a fixed mask ratio of 0.75, computed as mean squared error (MSE) over masked patches only. Pretraining used a custom FC-MAE wrapper and the encoder was instantiated as “convnextv2_base (timm)”. Images were patchified, and random masks were generated by shuffling patch indices and masking 75% of patch positions. For ConvNeXtV2, masking was applied in pixel space by zeroing out the pixels corresponding to masked patch regions before encoding (FC-MAE-style masking). A lightweight convolutional decoder (1 x 1 Conv > GELU > 1 x 1 Conv > GELU > 1 x 1 Conv) was used to predict reconstructed pixel patches, and the reconstruction loss was normalized by the number of masked patches. Pretraining was performed on RGB ECG images at 480 x 480 resolution. We applied a mild augmentation consisting of random resized cropping to the target resolution with a scale range of 0.8-1.0. The images used for ConvNeXtV2 pretraining were the same as those used for the ViT pretraining. Training used a custom PyTorch training loop. Pretraining was run for 160 epochs with a batch size of 64. Optimization used fused AdamW, and the learning rate followed a cosine schedule. Mixed-precision training was performed using autocast with bfloat16. Model checkpoints were saved at each epoch, and validation loss was evaluated and visualized to select the optimal pretrained weights, following the same procedure as in ViT pretraining.

**Self-supervised pretraining of MLP-Mixer (MAE)**

We performed self-supervised pretraining of an MLP-Mixer-B/16 backbone using a masked autoencoder (MAE) objective implemented in PyTorch with timm. The input consisted of square RGB ECG images (480 x 480 pixels). Images were patchified with a patch size of 16, and 75% of patch positions were randomly masked. Because the token-mixing MLP in MLP-Mixer depends on a fixed sequence length, we retained the full set of patch tokens in the encoder and replaced masked patch tokens with a learned mask token, rather than dropping masked tokens as in standard ViT-MAE. The model then reconstructed pixel values for each patch, and the reconstruction loss was computed as mean squared error (MSE) over masked patches only. The decoder was a lightweight per-token MLP (Linear > GELU repeated with a final Linear) that predicted 16 x 16 x 3 pixel values per patch.

For data augmentation during pretraining, we applied random resized cropping to the target resolution (scale 0.7 – 1.0; ratio 0.95 – 1.05), followed by random affine transformations (rotation, translation, scaling, and shear) and mild color jitter, and then converted images to tensors. When multiple image roots were used, datasets were concatenated and a weighted random sampler was applied to balance sampling across roots.

Pretraining was performed using a custom PyTorch training loop for up to 200 epochs, with a batch size of 128, fused AdamW optimization, and a cosine learning-rate schedule (base learning rate 3.27 x 10-4). Mixed-precision training was performed using autocast with bfloat16. Model checkpoints were saved at each epoch, and validation reconstruction loss was evaluated and visualized to select the optimal pretrained weights, following the same procedure as in ViT and ConvNeXtV2 pretraining.

**Finetuning**

Each pretrained model was fine-tuned using the training and validation datasets, first for 3-class classification and subsequently for binary classification. RandAugment without flip-like transformations, CutMix, and MixUp were applied during training where appropriate.[33-35] Hyperparameters were also selected by grid search using the internal validation dataset.

**Fine-tuning of ViT for 3-class classification using MAE-pretrained weights**

We fine-tuned a ViT image classifier for three-class prediction (Takotsubo syndrome, AMI, and non-Takotsubo/AMI).

**Input processing and augmentation**

All ECG images were loaded as RGB images and converted to tensors. Flip operations were explicitly excluded from data augmentation. For training, we used RandAugment with flip-like transforms removed, followed by RandomErasing. For validation and test sets, only tensor conversion and normalization were applied (no data augmentation). In addition, CutMix/Mixup data augmentation was implemented using the timm Mixup utility with label smoothing (0.1).

Model initialization from MAE-pretrained checkpoint

The ViT backbone was initialized from a previously self-supervised MAE pretraining checkpoint, and the classification head was replaced for three output classes. The classifier configuration was based on a ViT-Base patch-16 model and was updated to use an input size of 480 with increased regularization (hidden dropout probability 0.2; attention dropout probability 0.2).

**Training procedure**

Optimization and evaluation were conducted using the Hugging Face Transformers Trainer framework with Sharpness-Aware Minimization (SAM). Stochastic weight averaging (SWA) was applied via a callback. Training was performed for 20 epochs with a batch size of 64 and bfloat16 mixed-precision was enabled. Model checkpoints were saved at each epoch, and the checkpoint with lowest validation loss and no evidence of overfitting based on the training/validation loss curves was selected for downstream binary fine-tuning.

**Fine-tuning of ViT for 2-class classification (Takotsubo syndrome vs AMI)**

Following the 3-class fine-tuning, we further fine-tuned the ViT model for binary classification (Takotsubo syndrome vs AMI). The model was initialized from the best 3-class checkpoint, and the classifier head was replaced for two output classes. The same training/validation dataset was used, but only images labeled as Takotsubo syndrome or AMI were included. Input preprocessing and data augmentation were consistent with the 3-class setting. During training, RandAugment (with flip-like transforms removed) and CutMix/MixUp were applied; no data augmentation was applied to validation images.

Optimization and evaluation were performed using the Hugging Face Transformers Trainer framework with a custom SAM training step. For the binary task, we used focal loss with class-balanced weighting, with class weights proportional to the inverse square root of the class frequency in the training set. SWA was applied via a callback starting from 75% of total training steps.

Training was performed in two stages. First, the ViT backbone was frozen and only the classifier head was trained for three epochs using a cosine scheduling with a warmup ratio of 0.1. Second, all layers were unfrozen and training was continued for 15 epochs with differential learning rates applied to the backbone and head, again using a cosine schedule with warmup. Mixed-precision training was enabled with bfloat16. Checkpoints were saved and evaluated at each epoch, and the best model was selected using the same criterion as in the 3-class fine-tuning.

**Fine-tuning of ConvNeXtV2 for 3-class classification using FC-MAE-pretrained weights**

We fine-tuned a ConvNeXtV2-Base image classifier for three-class prediction (Takotsubo syndrome, AMI, and non-Takotsubo/AMI).

Input processing and data augmentation

ECG images were loaded as RGB images. For training, we used RandAugment with flip-like transforms removed, followed by tensor conversion, RandomErasing, and normalization. For validation, only tensor conversion and normalization were applied without data augmentation.

**Class balancing and Mixup/CutMix**

To mitigate class imbalance during training, we used a WeightedRandomSampler based on inverse class frequency to construct per-sample weights. CutMix/MixUp was implemented using the timm Mixup utility, producing soft targets during training.

**Model initialization from an FC-MAE checkpoint**

The ConvNeXtV2-Base backbone weights were initialized from the FC-MAE self-supervised checkpoint into the timm model.

**Training procedure**

Training was performed with a custom PyTorch training loop and epoch-wise checkpointing. During training, Mixup/CutMix soft targets were optimized using a soft-target cross-entropy loss, while validation used standard cross-entropy with integer labels. Model checkpoints were saved at each epoch, and the checkpoint with the lowest validation loss and no evidence of overfitting based on the training/validation loss curves was selected for downstream binary fine-tuning.

**Fine-tuning of ConvNeXtV2 for 2-class classification (Takotsubo vs AMI)**

Following the 3-class fine-tuning, we further fine-tuned ConvNeXtV2-Base for binary classification (Takotsubo syndrome vs AMI) using the same dataset structure, restricted to Takotsubo syndrome and AMI.

**Model initialization from the 3-class checkpoint**

The 2-class model was initialized from the 3-class fine-tuned checkpoint by transferring the backbone weights and adapting the classification head by selecting the Takotsubo and AMI class-specific weight vectors from the 3-class head and loading them into the 2-class head.

Input processing, data augmentation, and sampling

The same preprocessing/data augmentation strategy as in the 3-class setting was used, with no data augmentation for validation. Checkpoints were saved and evaluated at each epoch, and the best model was selected using the same criterion as in the 3-class fine-tuning.

**Fine-tuning of MLP-Mixer for 3-class classification using MAE-pretrained weights**

We fine-tuned an MLP-Mixer-B/16 image classifier (timm; mixer_b16_224) for three-class prediction (Takotsubo syndrome, AMI, and non-Takotsubo/AMI).

**Input processing and data augmentation**

ECG images were loaded as RGB images and resized to 480 x 480 pixels. Flip operations were excluded from data augmentation by using a RandAugment variant that removes flip-like transforms. For training, we applied RandAugmentNoFlip and a small random rotation, followed by tensor conversion and normalization. For validation, only resizing, tensor conversion, and normalization were applied without data augmentation.

**Class balancing and Mixup/CutMix**

To mitigate class imbalance, we used a WeightedRandomSampler with per-sample weights derived from inverse class frequency. Mixup/CutMix was implemented using timm Mixup, producing soft targets during training.

**Model initialization from an MAE checkpoint**

The model was created with an output head corresponding to the number of classes. Encoder weights were loaded from the MAE checkpoint by discarding decoder- and token-related parameters and loading the remaining shape-matched weights.

**Training procedures**

Training used a two-stage strategy: an initial head-only phase (backbone frozen) followed by full fine-tuning with the backbone unfrozen. During the head-only phase, Mixup/CutMix (and label smoothing) was disabled, and it was enabled in the subsequent full fine-tuning phase. Optimization used AdamW with separate parameter groups for backbone and head, and learning-rate scheduling used a cosine schedule with warmup for the full fine-tuning phase. SAM was applied during the full fine-tuning phase (not during head-only training), and SWA was also applied. Model checkpoints were saved each epoch, and the checkpoint with the most favorable validation-loss profile (lowest validation loss without evidence of overfitting based on training/validation loss curves) was selected for downstream binary fine-tuning.

**Fine-tuning of MLP-Mixer for 2-class classification (Takotsubo syndrome vs AMI)**

Following 3-class fine-tuning, we further fine-tuned MLP-Mixer-B/16 for binary classification (Takotsubo syndrome vs AMI) using the same dataset structure but restricting labels to Takotsubo syndrome and AMI.

**Model initialization from the 3-class checkpoint**

The 2-class model was initialized from the 3-class fine-tuning checkpoint. Backbone weights were transferred directly, and the classification head was adapted by selecting the Takotsubo and AMI class-specific weight and bias parameters from the 3-class head and loading them into the 2-class head.

**Input processing, data augmentation, and sampling**

Binary fine-tuning used the same preprocessing as 3-class fine-tuning (480 x 480 resizing; RGB; normalization), with flip-like transforms removed. RandAugmentNoFlip and small random rotation were applied during training, whereas validation used no augmentation. Class balancing again used WeightedRandomSampler, and Mixup/CutMix was implemented using timm Mixup to generate soft targets during training.

**Training and checkpoint selection**

Training was performed with epoch-wise checkpointing, and the best model was selected using the same validation-loss-based criterion as in the 3-class fine-tuning.

**The final ensemble model with TTA**

The above 3 fine-tuned models were designed to support TTA during inference. For each input ECG image, non-square images were first converted to a square image by either padding or center cropping and then resized to 480 x 480 pixels before model inference.

TTA was implemented by applying RandAugment with flip-like transforms removed (RandAugmentNoFlip) to generate multiple augmented views of the same 480 x 480 image and then averaging model outputs across views. Specifically, for ViT and MLP-Mixer, logits were computed for the original image and for N - 1 augmented images, and the mean logit vector was used as the model output for that ECG.

Model-specific TTA configurations were selected using the internal validation dataset only. ViT used 24 views with a RandAugment magnitude of 1, and MLP-Mixer used 32 views with a RandAugment magnitude of 3. For ConvNeXtV2, none of the evaluated TTA configurations improved performance; therefore, inference was performed using a single view (tta = 1), effectively without TTA.

For ensemble prediction, per-model logits were combined using equal weights (1:1:1), and a softmax function was applied to the ensemble logits to obtain the final output probability.

**Comparison of cross-architecture and same-architecture ensembles**

To examine the potential contribution of architectural diversity, we compared the final ensemble with same-architecture ensembles comprising three ViT, ConvNeXtV2, or MLP-Mixer models. For each architecture, candidates originated from distinct 3-class fine-tuning runs and were subsequently fine-tuned for binary classification. Three candidates were selected using low development-set validation loss and the absence of apparent overfitting in training and validation loss curves. Each same-architecture ensemble included the component used in the final ensemble. All ensembles used equal-weight logit averaging followed by softmax, the same external test images, and unchanged architecture-specific TTA settings. The same saved component predictions were reused across ensemble configurations, including the ViT predictions used in the primary analysis.

Paired differences in ROC AUC were calculated as the cross-architecture ensemble minus each same-architecture ensemble. Percentile 95% CIs were estimated using 5,000 patient-cluster bootstrap samples stratified by diagnosis. Each sample retained all images of sampled patients, including multiplicity, and was shared across ensembles. Trained models, predictions, and the AMI threshold of 0.5 remained fixed. Intervals were not adjusted for multiple comparisons; these comparisons were exploratory. A sensitivity analysis assigned each image weight 1/n, where n was the number of images from that patient. These intervals reflect patient-sampling uncertainty and do not include variability from model retraining or repeated stochastic TTA inference.

**Patient clustered evaluation of external model performance**

To account for within-patient correlation among ECG images, 95% CIs for accuracy, sensitivity, specificity, and ROC AUC were calculated using 5,000 patient-cluster bootstrap replicates, stratified by diagnosis. In each replicate, patients were sampled with replacement within each diagnostic group, preserving the original number of patients per group. All images from each selected patient were retained, including the multiplicity of patients selected more than once. Confidence limits were the 2.5th and 97.5th percentiles of the bootstrap distribution. The trained model, its predicted probabilities, and the AMI classification threshold of 0.5 were held fixed.

**Sensitivity analysis with equal patient weights**

To assess the influence of unequal image counts per patient, each image was assigned a weight equal to the reciprocal of that patient's number of evaluated images. Thus, each patient contributed a total weight of one. Weighted accuracy, sensitivity, specificity, and ROC AUC were calculated using these image weights; CIs used the same patient-cluster bootstrap draws as the primary evaluation. In each replicate, the image weight was multiplied by the number of times the corresponding patient was sampled. This analysis retained image-level predictions rather than combining images into a single patient-level diagnosis. Equal patient weighting changed the effective AMI proportion from 65.1% of images to 53.3% of patients; therefore, changes in overall accuracy also reflect the change in diagnostic composition (Supplementary Table S4).

**Selection and presentation of ECG images for the reader study**

Two sets of 50 ECG images were randomly sampled from the test set. Each set contained 25 AMI and 25 Takotsubo images. The set used in Phases 1 – 3 represented 33 patients (17 with AMI and 16 with Takotsubo syndrome), and the Phase 4 set represented 33 patients (18 with AMI and 15 with Takotsubo syndrome). Sixteen patients contributed to both sets, giving 50 distinct patients across the two sets. Multiple images from the same patient were included (Supplementary Table S6). Image order was randomized for each reader and retained across Phases 1 – 3; Phase 4 used a separately randomized order.

**Randomization in Phase 4 of the human diagnostic test**

For Phase 4, participants were assigned to one of three groups using computer-generated stratified block randomization with a 1:1:1 allocation ratio. Randomization was stratified according to cardiologist status and years of clinical experience, using four strata: cardiologists with >= 10 years of clinical experience, cardiologists with <10 years of clinical experience, non-cardiologists with >= 10 years of clinical experience, and non-cardiologists with <10 years of clinical experience. Within each stratum, randomization was performed using fixed blocks of size 3, with one allocation to each of the three groups in each block. The three groups were: (1) criteria-based reference sheet only; (2) reference sheet plus AI model predictions without Grad-CAM; and (3) reference sheet plus AI model predictions with Grad-CAM. The randomization table was generated before participant assignment using a dedicated computer script. At the start of Phase 4, each participant was assigned to the next available allocation slot within the corresponding stratum.

**Sample size estimation**

For the human diagnostic study, sample size estimation was based on participant-level accuracy. The key comparisons in Phases 1–3 were considered to be those between the unaided or reference sheet-assisted conditions and the AI-assisted condition. In the pilot data, mean accuracy differed by approximately 25 percentage points between Phase 3 and both Phases 1 and 2, resulting in a very small estimated sample size for these within-participant comparisons under a two-sided alpha level of 0.05 and 80% power. Because this estimate was driven by the large observed pilot effect and was considered unstable, the final sample size was determined based on the more conservative randomized comparison in Phase 4. Phase 4 was designed primarily to detect the difference between reference sheet alone and reference sheet plus AI assistance. In the pilot data, the effect size for the primary comparison between the non-AI-assisted group and the two AI-assisted groups combined was large, resulting in a small required sample size under a 1:1:1 allocation. However, for the secondary comparison evaluating the incremental effect of Grad-CAM, we anticipated that the difference between AI assistance with and without Grad-CAM could be smaller. Based on the pilot estimates, this comparison required approximately 16 participants per group, assuming a two-sided alpha level of 0.05 and 80% power. Because the participant-level accuracy analysis was planned using nonparametric tests, and to account for uncertainty in the pilot estimates and potential missing responses, the target sample size was increased to 20 participants per group. With 1:1:1 randomization to the three Phase 4 groups, the planned total sample size for the human diagnostic study was therefore 60 participants.

**Participant-level analyses of human diagnostic performance**

As a secondary participant-level analysis, accuracy was calculated for each participant within each phase or randomized group. Phases 1 – 3 were compared using the Friedman test, followed by paired Wilcoxon signed-rank tests for all three pairwise phase comparisons with Holm adjustment. Phase 4 groups were compared using the Kruskal-Wallis test. Mann-Whitney U tests compared the reference-sheet-only group with the two AI-assisted groups combined and the AI-assisted groups with vs without Grad-CAM, without multiplicity adjustment.

**Stratification adjusted sensitivity analyses for Phase 4**

Both specified Phase 4 comparisons were repeated with adjustment for the randomization stratification factors: cardiologist status and clinical experience categorized as < 10 or >= 10 years. Separate binomial GLMMs with a logit link included random intercepts for reader and ECG image case. The primary model compared both AI-assisted groups combined with the reference-sheet-only group among all 70 readers; the secondary model compared AI assistance with vs without Grad-CAM among the 48 AI-assisted readers. The two adjustment factors were included as additive fixed effects. Models were fitted by maximum likelihood using the Laplace approximation and the bobyqa optimizer, with a maximum of 200,000 function evaluations, in R 4.6.1 with lme4 2.0.1. ORs and Wald 95% CIs were obtained from the treatment coefficient and its standard error. Two-sided Wald P values were calculated without additional multiplicity adjustment for these two specified comparisons. The corresponding unadjusted models were refitted using the same input data. No responses were excluded, and no missing covariates required imputation. All four models converged without singular fits (Supplementary Table S7).

### Study protocol and registration

The study protocol is available from the corresponding author upon request. The study was not registered.

### Patient and public involvement

Patients and members of the public were not involved in the design of this study.

**Data, code, and model availability**

The institutional ECG data underlying this article cannot be shared publicly to protect patient privacy and will be shared on reasonable request to the corresponding author, subject to institutional approval. The public ECG databases used for pretraining are available from PhysioNet.11-14 The training code, trained models, and example inference code are available on GitHub (https://github.com/ekagawa007/takotsubo_ami_ai_ecg). Software version 0.4.0 is archived at https://doi.org/10.5281/zenodo.23763944. Trained model weights version 0.2.0 are archived at https://doi.org/10.5281/zenodo.23666262 and are also distributed at https://huggingface.co/ekagawa/takotsubo-ami-ai-ecg.

**Supplementary Table S1. External validation performance of AI models according to pretraining strategy and TTA**

| Component model (backbone) | Pretraining | TTA | Accuracy | Precision | Recall (sensitivity) | Specificity | F1 score | ROC AUC | PR AUC | ECE | Brier score | BSS |
|---|---|---|---|---|---|---|---|---|---|---|---|---|
| ViT | - | - | 0.654 | 0.748 | 0.705 | 0.558 | 0.726 | 0.719 | 0.850 | 0.103 | 0.213 | 0.063 |
| ViT | - | + | 0.648 | 0.735 | 0.717 | 0.519 | 0.726 | 0.722 | 0.853 | 0.114 | 0.219 | 0.037 |
| ViT | ImageNet | - | 0.809 | 0.866 | 0.836 | 0.758 | 0.851 | 0.887 | 0.943 | 0.053 | 0.133 | 0.414 |
| ViT | ImageNet | + | 0.821 | 0.870 | 0.852 | 0.763 | 0.861 | 0.902 | 0.951 | 0.057 | 0.126 | 0.447 |
| ViT | SSL | - | 0.875 | 0.909 | 0.898 | 0.832 | 0.903 | 0.947 | 0.973 | 0.095 | 0.095 | 0.580 |
| ViT | SSL | + | 0.882 | 0.908 | 0.912 | 0.828 | 0.910 | 0.951 | 0.974 | 0.094 | 0.094 | 0.584 |
| ConvNeXtV2* | - | - | 0.707 | 0.783 | 0.761 | 0.607 | 0.772 | 0.767 | 0.867 | 0.112 | 0.203 | 0.108 |
| ConvNeXtV2 | ImageNet | - | 0.896 | 0.910 | 0.933 | 0.827 | 0.921 | 0.942 | 0.948 | 0.018 | 0.080 | 0.649 |
| ConvNeXtV2 | SSL | - | 0.899 | 0.892 | 0.962 | 0.782 | 0.925 | 0.963 | 0.977 | 0.039 | 0.076 | 0.666 |
| MLP-Mixer† | - | - | 0.732 | 0.791 | 0.799 | 0.608 | 0.795 | 0.788 | 0.862 | 0.150 | 0.204 | 0.103 |
| MLP-Mixer | - | + | 0.732 | 0.805 | 0.776 | 0.650 | 0.790 | 0.807 | 0.892 | 0.075 | 0.177 | 0.219 |
| MLP-Mixer | SSL | - | 0.900 | 0.916 | 0.931 | 0.841 | 0.924 | 0.955 | 0.977 | 0.083 | 0.091 | 0.602 |

| | | | | | | | | | | | | |
|---|---|---|---|---|---|---|---|---|---|---|---|---|
| MLP-Mixer | SSL | + | 0.907 | 0.919 | 0.939 | 0.847 | 0.929 | 0.959 | 0.978 | 0.069 | 0.081 | 0.642 |
| Final ensemble model | | | 0.914 | 0.920 | 0.950 | 0.846 | 0.935 | 0.970 | 0.984 | 0.025 | 0.065 | 0.712 |

AI indicates artificial intelligence; SSL, self-supervised learning; TTA, test-time augmentation; ROC, receiver-operating-characteristics; AUC, area-under-curve; PR, precision-recall; ECE, expected calibration error; and BSS, Brier skill score.

*No favorable TTA hyperparameters were identified during TTA tuning for the ConvNeXtV2 backbone on the development dataset, and TTA was not applied to the ConvNeXtV2 model in the final inference pipeline.

†ImageNet-pretrained MLP-Mixer weights compatible with 480 x 480-pixel input images were not available.

‡SSL was performed using publicly available ECG datasets and the development dataset from the present study.

**Supplementary Table S2. Comparison of cross-architecture and same-architecture ensembles**

**A. Ensemble performance**

| Ensemble | ROC AUC (95% CI) | Accuracy | Sensitivity | Specificity | Brier score |
|---|---|---|---|---|---|
| ViT + ConvNeXtV2 + MLP-Mixer | 0.970 (0.938 – 0.992) | 0.914 | 0.950 | 0.846 | 0.065 |
| ViT only | 0.952 (0.913 – 0.981) | 0.879 | 0.907 | 0.826 | 0.101 |
| ConvNeXtV2 only | 0.965 (0.930 – 0.988) | 0.898 | 0.948 | 0.804 | 0.074 |
| MLP-Mixer only | 0.959 (0.920 – 0.987) | 0.906 | 0.939 | 0.845 | 0.082 |

**Supplementary Table S2. Continued**

**B. Paired differences relative to same-architecture ensembles**

| Same-architecture comparator | Difference in ROC AUC (95% CI) | Difference in accuracy (95% CI) | Patient-equal-weighted difference in ROC AUC (95% CI) |
|---|---|---|---|
| ViT only | 0.0180 (0.0045 – 0.0356) | 0.0347 (0.0077 – 0.0644) | 0.0242 (0.0066 – 0.0470) |
| ConvNeXtV2 only | 0.0050 (-0.0071 – 0.0196) | 0.0156 (-0.0063 – 0.0411) | 0.0102 (-0.0040 – 0.0278) |
| MLP-Mixer only | 0.0112 (0.0028 – 0.0221) | 0.0073 (0.0002 – 0.0142) | 0.0106 (0.0024 – 0.0210) |

All analyses used the same 8,883 images from 75 patients (40 AMI and 35 Takotsubo syndrome). Each ensemble comprised three models and used equal-weight logit averaging followed by softmax. AMI was the positive class; the classification threshold was 0.5.

Differences are the cross-architecture ensemble minus the comparator. CIs are percentile intervals from 5,000 diagnosis-stratified patient-cluster bootstrap samples shared across ensembles. They were not adjusted for multiple comparisons; comparisons were exploratory. Patient-equal weighting assigns each image weight 1/n, where n is that patient’s image count. Models and predictions were fixed.

AMI indicates acute myocardial infarction; AUC, area-under-curve; CI, confidence interval; and ROC, receiver-operating-characteristics.

**Supplementary Table S2. Continued**

**C. Performance of the nine component models**

| Component model | ROC AUC | Accuracy | Precision | Sensitivity | Specificity | F1 score | Brier score |
|---|---|---|---|---|---|---|---|
| ViT 1* | 0.951 | 0.882 | 0.908 | 0.912 | 0.828 | 0.910 | 0.094 |
| ViT 2 | 0.949 | 0.875 | 0.905 | 0.904 | 0.822 | 0.904 | 0.106 |
| ViT 3 | 0.947 | 0.876 | 0.902 | 0.909 | 0.816 | 0.905 | 0.108 |
| ConvNeXtV2 1* | 0.963 | 0.899 | 0.892 | 0.962 | 0.782 | 0.925 | 0.076 |
| ConvNeXtV2 2 | 0.961 | 0.897 | 0.912 | 0.931 | 0.833 | 0.921 | 0.078 |
| ConvNeXtV2 3 | 0.958 | 0.897 | 0.896 | 0.952 | 0.794 | 0.923 | 0.078 |
| MLP-Mixer 1* | 0.959 | 0.907 | 0.919 | 0.939 | 0.847 | 0.929 | 0.081 |
| MLP-Mixer 2 | 0.959 | 0.907 | 0.919 | 0.940 | 0.845 | 0.929 | 0.082 |
| MLP-Mixer 3 | 0.959 | 0.907 | 0.918 | 0.940 | 0.845 | 0.929 | 0.082 |

*Component included in the final cross-architecture ensemble. Within each architecture, models originated from distinct 3-class fine-tuning runs and were selected using development-set validation loss and assessment of overfitting. Numbers identify models and do not denote test-set performance ranks.

Architecture-specific TTA settings were unchanged: 24 views with magnitude 1 for ViT, 32 views with magnitude 3 for MLP-Mixer, and no TTA for ConvNeXtV2. AMI was the positive class for precision, sensitivity, and F1 score. All values in panel C are image-weighted.

TTA indicates test-time augmentation.

**Supplementary Table S3. External validation performance of the AI models using images derived from first recorded ECGs**

| | Accuracy | Precision | Recall (sensitivity) | Specificity | F1 score | ROC AUC | PR AUC | ECE | Brier score | BSS |
|---|---|---|---|---|---|---|---|---|---|---|
| Component model (backbone) | | | | | | | | | | |
| ViT | 0.855 | 0.840 | 0.902 | 0.802 | 0.869 | 0.928 | 0.940 | 0.085 | 0.115 | 0.538 |
| ConvNeXtV2 | 0.861 | 0.813 | 0.962 | 0.746 | 0.881 | 0.958 | 0.962 | 0.094 | 0.102 | 0.588 |
| MLP-Mixer | 0.866 | 0.828 | 0.946 | 0.774 | 0.883 | 0.945 | 0.958 | 0.128 | 0.122 | 0.511 |
| Final ensemble model | 0.864 | 0.822 | 0.951 | 0.763 | 0.882 | 0.958 | 0.965 | 0.093 | 0.097 | 0.609 |

Performance was evaluated at the image level using 836 images derived from the first recorded ECGs.

AI indicates artificial intelligence; ROC, receiver-operating-characteristics; AUC, area-under-curve; PR, precision-recall; ECE, expected calibration error; and BSS, Brier skill score.

**Supplementary Table S4. External validation with patient-level sensitivity analysis**

| | Image-weighted estimate (95% CI) | Patient-equal-weighted estimate (95% CI) |
|---|---|---|
| Accuracy | 0.914 (0.866 – 0.954) | 0.903 (0.854 – 0.945) |
| Sensitivity | 0.950 (0.894 – 0.988) | 0.949 (0.896 – 0.987) |
| Specificity | 0.846 (0.746 – 0.928) | 0.850 (0.762 – 0.925) |
| ROC AUC | 0.970 (0.938 – 0.992) | 0.968 (0.937 – 0.991) |

Both analyses used the same 8,883 images from 75 patients (40 AMI and 35 Takotsubo syndrome). CIs are percentile intervals from 5,000 bootstrap samples of patients, stratified by diagnosis. The trained model and AMI threshold of 0.5 were fixed.

Patient-equal weighting assigns each image weight 1/n, where n is that patient's image count, so each patient contributes total weight one. These are weighted image-level predictions, not a single prediction per patient. The effective AMI proportion is 65.1% in the image-weighted analysis and 53.3% with equal-patient weights.

AMI indicates acute myocardial infarction; AUC, area-under-curve; CI, confidence interval; ROC, receiver-operating-characteristics.

**Supplementary Table S5. Characteristics of the human test participants**

| | All | Group 0 (reference sheet) | Group 1 (AI without Grad-CAM) | Group 2 (AI with Grad-CAM) | P |
|---|---|---|---|---|---|
| Participant, n | 70 | 22 | 23 | 25 | |
| Physicians, n | 49 (70) | 13 (59) | 19 (83) | 17 (68) | 0.22 |
| Cardiologist, n | 36 (51) | 12 (55) | 12 (52) | 12 (48) | 0.90 |
| Non-cardiologist physician, n | 13 (19) | 1 (5) | 7 (30) | 5 (20) | |
| Non-physician, n | 21 (30) | 9 (41) | 4 (17) | 8 (32) | |
| Experience, y | 11 [6 – 19] | 12 [5 – 18] | 10 [6 – 14] | 11 [8 – 20] | 0.68 |
| < 10 years, n | 31 (44) | 10 (45) | 11 (48) | 10 (40) | 0.85 |
| >= 10 years, n | 39 (56) | 12 (55) | 12 (52) | 15 (60) | 0.85 |

Data were presented as n (%) or median [interquartile range]. Randomization was stratified by cardiologist status and experience (cut-off 10 years).

AI indicates artificial intelligence; and Grad-CAM, gradient-weighted class activation mapping.

**Supplementary Table S6. Composition of the ECG image sets used in the reader study**

| | Phases 1 – 3 | Phase 4 |
|---|---|---|
| Presented image files, n | 50 | 50 |
| AMI image files, n | 25 | 25 |
| Takotsubo syndrome image files, n | 25 | 25 |
| Patients, n | 33 | 33 |
| AMI patients, n | 17 | 18 |
| Takotsubo syndrome patients, n | 16 | 15 |
| Patients with multiple presented images, n | 8 | 12 |
| Images per patient, median (range) | 1 (1 – 4) | 1 (1 – 4) |
| Original ECG recordings, n | 50 | 50 |

Images were randomly sampled from the test set. Sixteen patients (9 AMI and 7 Takotsubo syndrome) contributed to both sets. There was no identical source ECG between sets. Phases 1 – 3 used the same image set.

AMI indicates acute myocardial infarction; and ECG, electrocardiogram.

**Supplementary Table S7. Phase 4 sensitivity analyses adjusted for the randomization stratification factors**

| | Unadjusted OR (95% CI) | P value | Adjusted OR (95% CI) | P value |
|---|---|---|---|---|
| AI support vs reference sheet only | 4.28 (3.08 – 5.95) | < 0.001 | 4.31 (3.11 – 5.96) | < 0.001 |
| AI + Grad-CAM vs AI without Grad-CAM | 1.53 (1.04 – 2.25) | 0.033 | 1.54 (1.07 – 2.21) | 0.019 |

The AI vs reference-sheet only comparison included 70 readers and 3,500 responses. The Grad-CAM comparison included the 48 AI-assisted readers and 2,400 responses. All models used 50 ECG image cases and random intercepts for reader and case.

Adjusted models include cardiologist status and clinical experience (< 10 vs >= 10 years) as additive fixed effects. ORs, Wald 95% intervals, and two-sided Wald P values were estimated using binomial GLMMs with a logit link. No additional multiplicity adjustment was applied to these two specified comparisons. All 4 models converged without singular fits.

AI indicates artificial intelligence; CI, confidence interval; ECG, electrocardiogram; Grad-CAM, gradient-weighted class activation mapping; and OR indicates odds ratio.

**Supplementary Table S8. Participant counts by development-set partition**

| Partition | Takotsubo syndrome | AMI | Healthy volunteers | Total |
|---|---|---|---|---|
| Training | 235 | 243 | 155 | 633 |
| Validation | 55 | 58 | 36 | 149 |
| Pretraining-validation | 18 | 20 | 12 | 50 |
| Total | 308 | 321 | 203 | 832 |

Counts are participants. Partitions were mutually exclusive at the participant level. The pretraining-validation partition was used for self-supervised pretraining validation.

AMI indicates acute myocardial infarction; and ECG, electrocardiogram.

**Supplementary Figure S1. Criteria-based electrocardiographic reference sheet for differentiating Takotsubo syndrome from acute myocardial infarction.**

This reference sheet, developed on the basis of previously reported electrocardiographic criteria, was provided to participants in Phases 2–4 of the human diagnostic test.

AMI indicates acute myocardial infarction; NSTEMI, non-ST-segment elevation myocardial infarction; NSTE, non-ST-segment elevation; and STEMI, ST-segment elevation myocardial infarction.

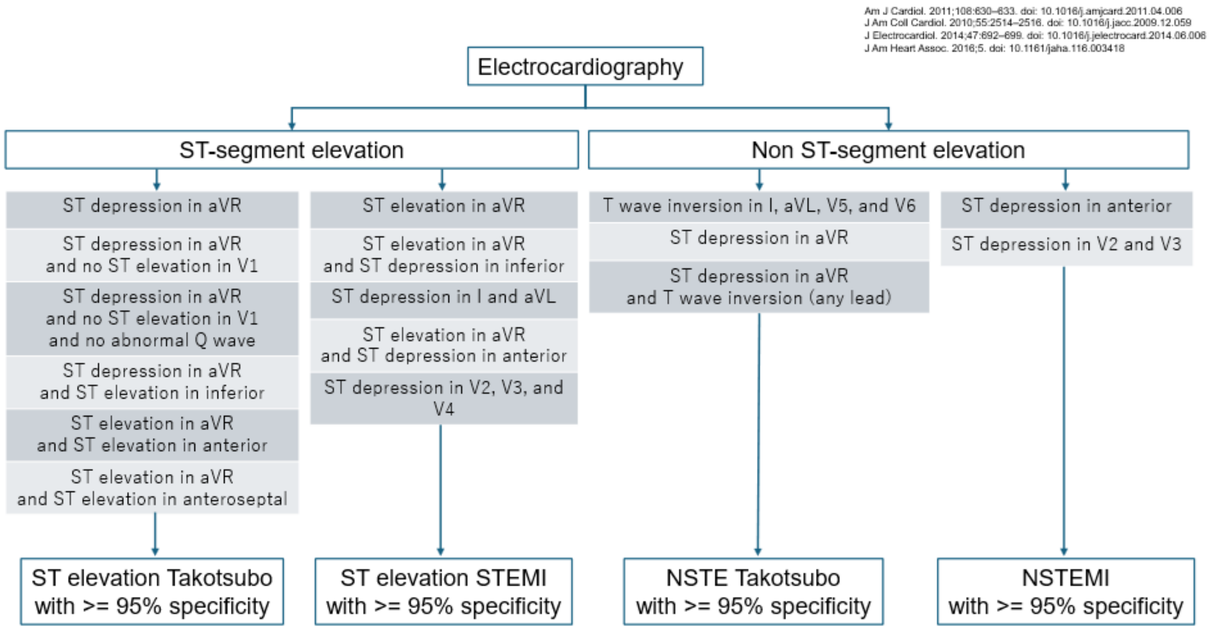


STEMI, ST-segment elevation myocardial infarction; NSTE, non-ST-segment elevation; NSTEMI, non-ST-segment elevation myocardial infarction.

**Supplementary Figure S2. Representative example of AI predictions for Takotsubo syndrome and AMI.**

Representative electrocardiograms from a patient with Takotsubo syndrome (A) and a patient with AMI (B). Each panel shows the original ECG, the resized model input (480 x 480 pixels), and the predicted probabilities for Takotsubo syndrome and AMI from the three individual models (ViT, ConvNeXtV2, and MLP-Mixer) and their ensemble. Visualizations include a ViT attention map, Grad-CAM heatmaps for ViT and ConvNeXtV2, and the corresponding Grad-CAM overlays on the input ECG. Grad-CAM heatmaps are shown for the predicted class: Takotsubo syndrome in panel A and AMI in panel B.

AI indicates artificial intelligence; AMI, acute myocardial infarction; ECG, electrocardiogram; and Grad-CAM, gradient-weighted class activation mapping.

**A)**

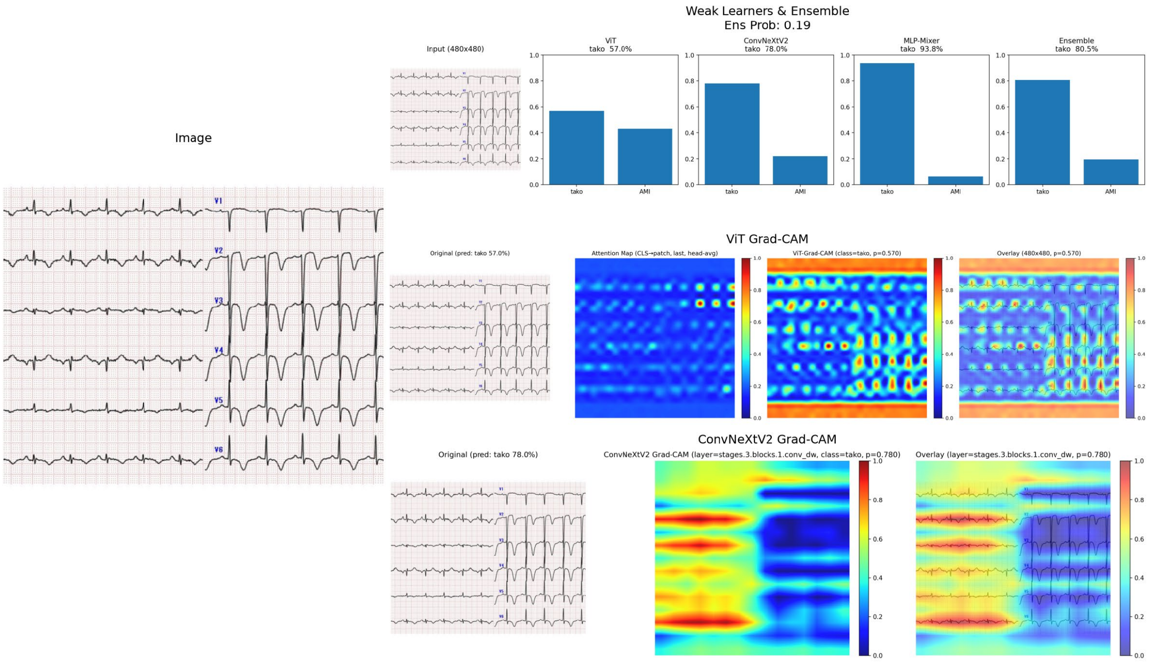
Weak Learners & Ensemble
Ens Prob: 0.19
Input (480x480)
ViT
tako 57.0%
ConvNeXtV2
tako 78.0%
MLP-Mixer
tako 93.8%
Ensemble
tako 80.5%
tako
AMI
Image
V1
V2
V3
V4
V5
V6
ViT Grad-CAM
Original (pred: tako 57.0%)
Attention Map (CLS→patch, last, head-avg)
ViT-Grad-CAM (class=tako, p=0.570)
Overlay (480x480, p=0.570)
ConvNeXtV2 Grad-CAM
Original (pred: tako 78.0%)
ConvNeXtV2 Grad-CAM (layer=stages.3.blocks.1.conv_dw, class=tako, p=0.780)
Overlay (layer=stages.3.blocks.1.conv_dw, p=0.780)

**B)**

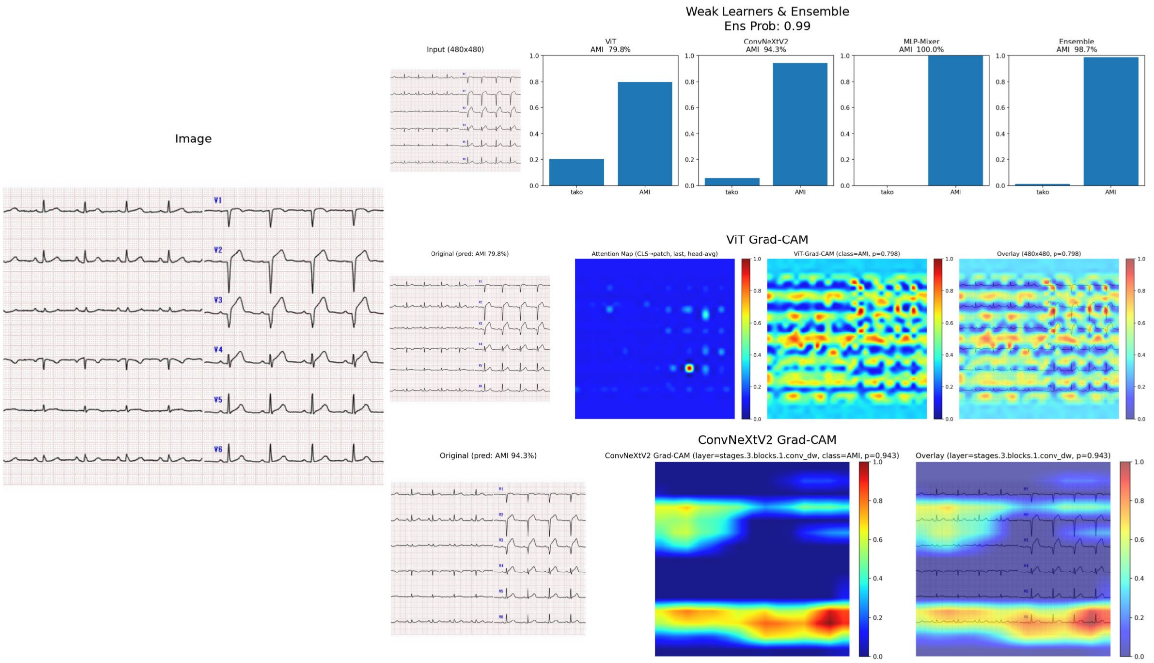
Weak Learners & Ensemble
Ens Prob: 0.99
Input (480x480)
ViT
AMI 79.8%
ConvNeXtV2
AMI 94.3%
MLP-Mixer
AMI 100.0%
Ensemble
AMI 98.7%
tako
AMI
Image
V1
V2
V3
V4
V5
V6
ViT Grad-CAM
Original (pred: AMI 79.8%)
Attention Map (CLS→patch, last, head-avg)
ViT-Grad-CAM (class=AMI, p=0.798)
Overlay (480x480, p=0.798)
ConvNeXtV2 Grad-CAM
Original (pred: AMI 94.3%)
ConvNeXtV2 Grad-CAM (layer=stages.3.blocks.1.conv_dw, class=AMI, p=0.943)
Overlay (layer=stages.3.blocks.1.conv_dw, p=0.943)

**Supplementary Figure S3. Performance of the final ensemble model using images derived from each patient's first recorded ECG.**

ROC curve (A) and PR curve (B) of the final ensemble model for predicting AMI. The confusion matrix (C) shows the number of ECG images in each true and predicted diagnostic category. Performance was evaluated at the image level using 836 images.

AMI indicates acute myocardial infarction; ECG, electrocardiogram; PR, precision–recall; and ROC, receiver-operating characteristic.

A)

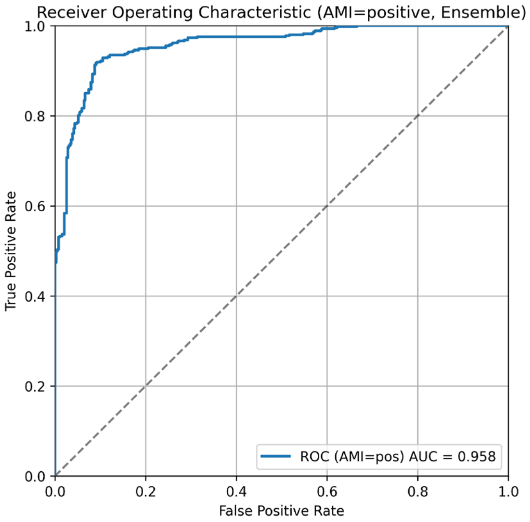
Receiver Operating Characteristic (AMI=positive, Ensemble)
True Positive Rate
False Positive Rate
0.0
0.2
0.4
0.6
0.8
1.0
ROC (AMI=pos) AUC = 0.958

B)

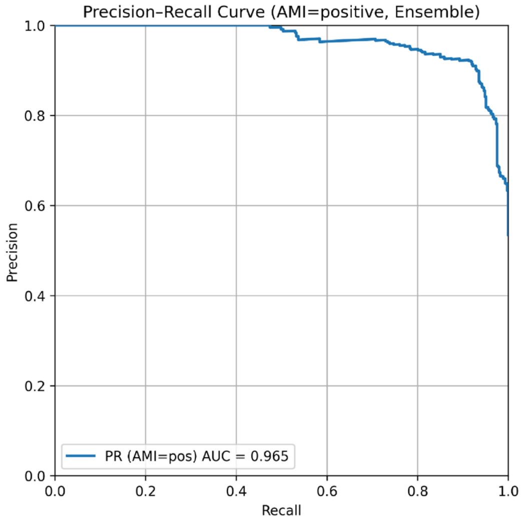
Precision–Recall Curve (AMI=positive, Ensemble)
Precision
Recall
0.0
0.2
0.4
0.6
0.8
1.0
PR (AMI=pos) AUC = 0.965

C)

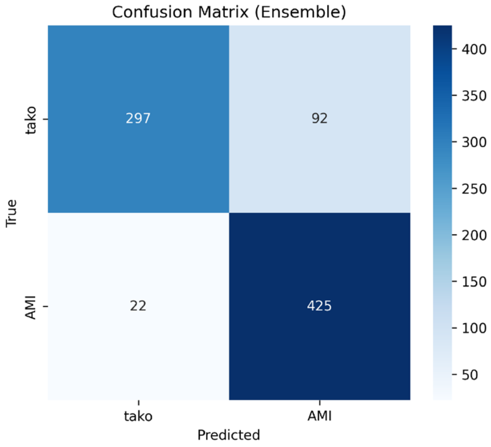
Confusion Matrix (Ensemble)
True
Predicted
tako
AMI
297
92
22
425
400
350
300
250
200
150
100
50

**Supplementary Figure S4. Phase 4 pairwise group odds ratios.**

Odds ratio (95% confidence interval) for a correct diagnosis from a generalized linear mixed model with random intercepts for participants and ECG case. For the three pairwise group comparisons, P values were adjusted using the Holm method, and 95% CIs for the ORs were adjusted using the Bonferroni method. Compared with the reference sheet alone (group 0), accuracy was higher with AI assistance without Grad-CAM (group 1) and with AI assistance + Grad-CAM (group 2) after Holm adjustment.

AI indicates artificial intelligence; and Grad-CAM, gradient-weighted class activation mapping.

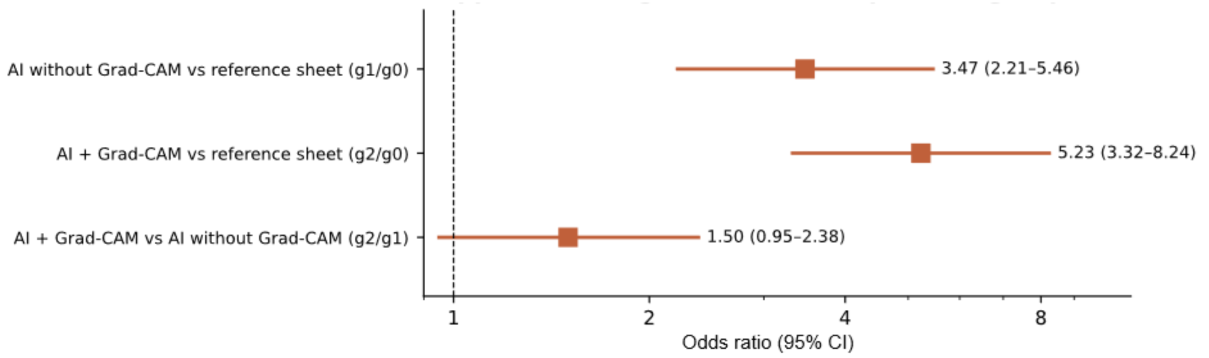

**Supplementary Figure S5. Accuracy by subgroup across Phases 1 – 3.**

Participant-level accuracy in Phase 1 (unaided), Phase 2 (reference sheet), and Phase 3 (reference sheet + AI assistance), shown separately by (A) years of clinical experience and (B) cardiologist status. Mean unaided accuracy was numerically higher among cardiologists and more experienced readers, although only the association with cardiologist status was statistically significant. With AI assistance (Phase 3), the subgroup lines crossed: less experienced readers and non-cardiologists achieved higher accuracy than more experienced readers and cardiologists, indicating that the incremental benefit of AI assistance was greatest among readers with less prior expertise. Corresponding within-phase ORs are shown in Supplementary Figure S6.

AI indicates artificial intelligence; and SD, standard deviation.

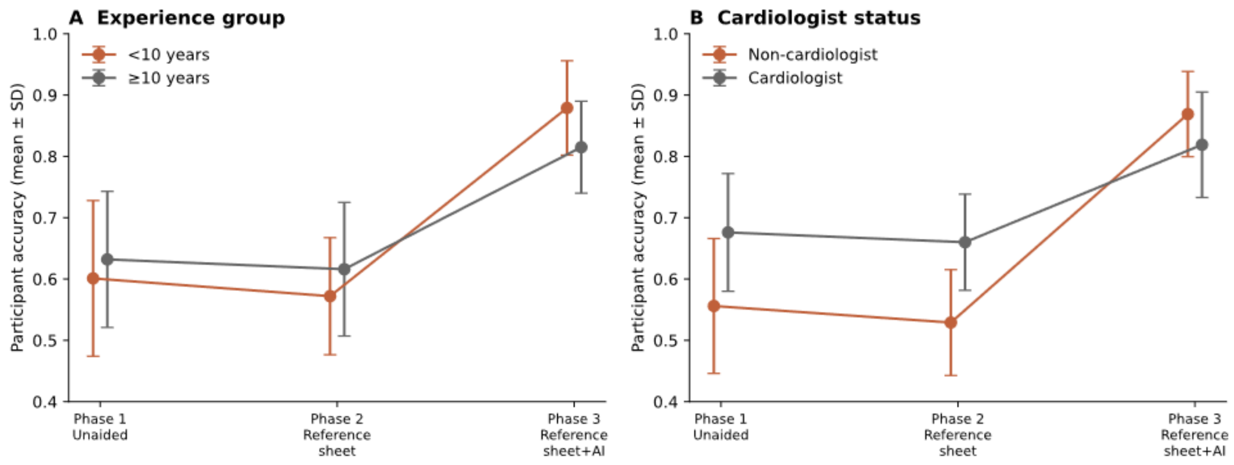

**Supplementary Figure S6. Within-phase subgroup odds ratios (Phases 1 – 3).**

ORs estimated using GLMMs with random intercepts for participant and ECG case, comparing readers with >= 10 vs < 10 years of clinical experience and cardiologists vs non-cardiologists within each phase. Without AI assistance, cardiologists outperformed non-cardiologists, whereas experience alone showed no significant effect. Under AI assistance (Phase 3), both associations reversed.

AI indicates artificial intelligence; CI, confidence interval; GLMM, generalized linear mixed model; and OR, odds ratio.

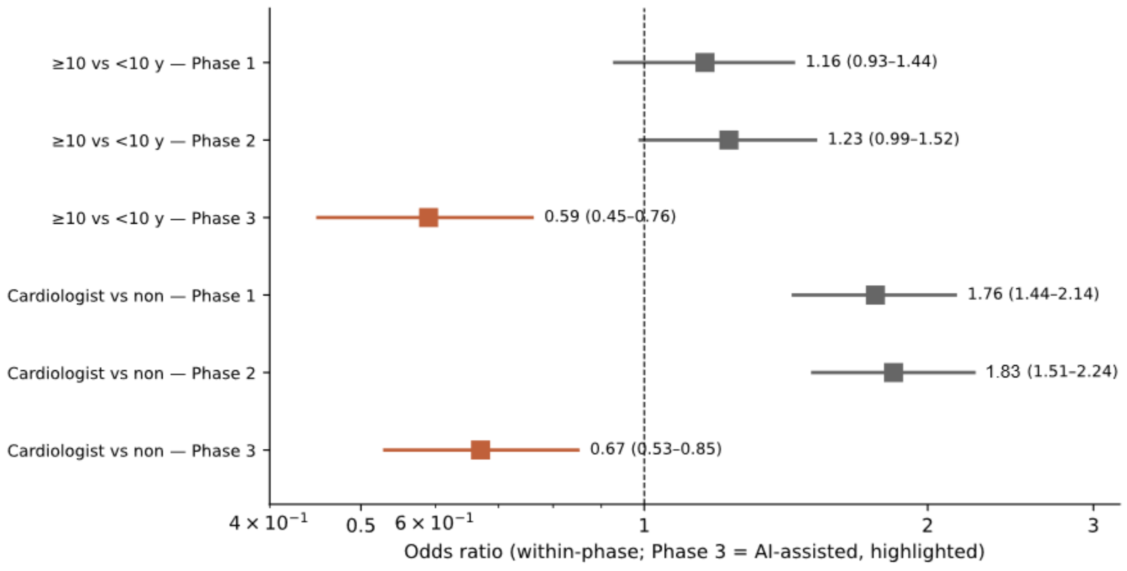

### Supplementary TRIPOD+AI checklist

TRIPOD+AI

Version: 11-January-2024

M = Manuscript; S = Supplement. T1, T2, T3 = separate Tables 1, 2, 3 (pages 1 - 3, 1 - 2, 1 - 2, respectively).
F2D = Figure 2D. NR = not reported. NA = not applicable. An asterisk indicates partial reporting.

| Section/Topic | Item | Development / evaluation[1] | Checklist item | Reported on page |
|---|---|---|---|---|
| **TITLE** | | | | |
| *Title* | 1 | D;E | Identify the study as developing or evaluating the performance of a multivariable prediction model, the target population, and the outcome to be predicted | M1* |
| **ABSTRACT** | | | | |
| *Abstract* | 2 | D;E | See TRIPOD+AI for Abstracts checklist | M3-4* |
| **INTRODUCTION** | | | | |
| *Background* | 3a | D;E | Explain the healthcare context (including whether diagnostic or prognostic) and rationale for developing or evaluating the prediction model, including references to existing models | M6-7, 16 |
| | 3b | D;E | Describe the target population and the intended purpose of the prediction model in the context of the care pathway, including its intended users (e.g., healthcare professionals, patients, public) | M7, 15-16 |
| | 3c | D;E | Describe any known health inequalities between sociodemographic groups | NR |
| *Objectives* | 4 | D;E | Specify the study objectives, including whether the study describes the development or validation of a prediction model (or both) | M7 |
| **METHODS** | | | | |
| *Data* | 5a | D;E | Describe the sources of data separately for the development and evaluation datasets (e.g., randomised trial, cohort, routine care or registry data), the rationale for using these data, and representativeness of the data | M7 - 9, S2-3 |
| | 5b | D;E | Specify the dates of the collected participant data, including start and end of participant accrual; and, if applicable, end of follow-up | M7-8* |
| *Participants* | 6a | D;E | Specify key elements of the study setting (e.g., primary care, secondary care, general population) including the number and location of centres | M7-9 |
| | 6b | D;E | Describe the eligibility criteria for study participants | M7-8, S3 |
| | 6c | D;E | Give details of any treatments received, and how they were handled during model development or evaluation, if relevant | M7-8, 12-13 |
| *Data preparation* | 7 | D;E | Describe any data pre-processing and quality checking, including whether this was similar across relevant sociodemographic groups | S2-3, 12-13* |
| *Outcome* | 8a | D;E | Clearly define the outcome that is being predicted and the time horizon, including how and when assessed, the rationale for choosing this outcome, and whether the method of outcome assessment is consistent across sociodemographic groups | M7-8, S3 |
| | 8b | D;E | If outcome assessment requires subjective interpretation, describe the qualifications and demographic characteristics of the outcome assessors | S3* |
| | 8c | D;E | Report any actions to blind assessment of the outcome to be predicted | NR |
| *Predictors* | 9a | D | Describe the choice of initial predictors (e.g., literature, previous models, all available predictors) and any pre-selection of predictors before model building | M9-10, 16-17 |
| | 9b | D;E | Clearly define all predictors, including how and when they were measured (and any actions to blind assessment of predictors for the outcome and other predictors) | M7-8, S2, 12-13* |
| | 9c | D;E | If predictor measurement requires subjective interpretation, describe the qualifications and demographic characteristics of the predictor assessors | NA |
| *Sample size* | 10 | D;E | Explain how the study size was arrived at (separately for development and evaluation), and justify that the study size was sufficient to answer the research question. Include details of any sample size calculation | M7-8, S16-17* |
| *Missing data* | 11 | D;E | Describe how missing data were handled. Provide reasons for omitting any data | S3, 18* |
| *Analytical methods* | 12a | D | Describe how the data were used (e.g., for development and evaluation of model performance) in the analysis, including whether the data were partitioned, considering any sample size requirements | M8-9, S30 |
| | 12b | D | Depending on the type of model, describe how predictors were handled in the analyses (functional form, rescaling, transformation, or any standardisation). | S7, 9-13 |
| | 12c | D | Specify the type of model, rationale[2], all model-building steps, including any hyperparameter tuning, and method for internal validation | S3-14* |
| | 12d | D;E | Describe if and how any heterogeneity in estimates of model parameter values and model performance was handled and quantified across clusters (e.g., hospitals, countries). See TRIPOD-Cluster for additional considerations[3] | NA |
| | 12e | D;E | Specify all measures and plots used (and their rationale) to evaluate model performance (e.g., discrimination, calibration, clinical utility) and, if relevant, to compare multiple models | M10, S14-15 |
| | 12f | E | Describe any model updating (e.g., recalibration) arising from the model evaluation, either overall or for particular sociodemographic groups or settings | M10 |
| | 12g | E | For model evaluation, describe how the model predictions were calculated (e.g., formula, code, object, application programming interface) | S12-13 |
| *Class imbalance* | 13 | D;E | If class imbalance methods were used, state why and how this was done, and any subsequent methods to recalibrate the model or the model predictions | S8-9, 11-12 |
| *Fairness* | 14 | D;E | Describe any approaches that were used to address model fairness and their rationale | NR |
| *Model output* | 15 | D | Specify the output of the prediction model (e.g., probabilities, classification). Provide details and rationale for any classification and how the thresholds were identified | M10* |

[1] D=items relevant only to the development of a prediction model; E=items relating solely to the evaluation of a prediction model; D;E=items applicable to both the development and evaluation of a prediction model
[2] Separately for all model building approaches.
[3] TRIPOD-Cluster is a checklist of reporting recommendations for studies developing or validating models that explicitly account for clustering or explore heterogeneity in model performance (eg, at different hospitals or centres). Debray et al, BMJ 2023; 380: e071018 [DOI: 10.1136/bmj-2022-071018]

TRIPOD+AI

Version: 11-January-2024

| | | | | |
|---|---|---|---|---|
| *Training versus evaluation* | 16 | D;E | Identify any differences between the development and evaluation data in healthcare setting, eligibility criteria, outcome, and predictors | M7-9, T1 |
| *Ethical approval* | 17 | D;E | Name the institutional research board or ethics committee that approved the study and describe the participant-informed consent or the ethics committee waiver of informed consent | M8-9 |
| **OPEN SCIENCE** | | | | |
| *Funding* | 18a | D;E | Give the source of funding and the role of the funders for the present study | M22 |
| *Conflicts of interest* | 18b | D;E | Declare any conflicts of interest and financial disclosures for all authors | M22 |
| *Protocol* | 18c | D;E | Indicate where the study protocol can be accessed or state that a protocol was not prepared | S18 |
| *Registration* | 18d | D;E | Provide registration information for the study, including register name and registration number, or state that the study was not registered | S18 |
| *Data sharing* | 18e | D;E | Provide details of the availability of the study data | S19 |
| *Code sharing* | 18f | D;E | Provide details of the availability of the analytical code[4] | S19 |
| **PATIENT & PUBLIC INVOLVEMENT** | | | | |
| *Patient & Public Involvement* | 19 | D;E | Provide details of any patient and public involvement during the design, conduct, reporting, interpretation, or dissemination of the study or state no involvement. | S18* |
| **RESULTS** | | | | |
| *Participants* | 20a | D;E | Describe the flow of participants through the study, including the number of participants with and without the outcome and, if applicable, a summary of the follow-up time. A diagram may be helpful. | M30-31, S30, T1* |
| | 20b | D;E | Report the characteristics overall and, where applicable, for each data source or setting, including the key dates, key predictors (including demographics), treatments received, sample size, number of outcome events, follow-up time, and amount of missing data. A table may be helpful. Report any differences across key demographic groups. | S30, T1 |
| | 20c | E | For model evaluation, show a comparison with the development data of the distribution of important predictors (demographics, predictors, and outcome). | T1 |
| *Model development* | 21 | D;E | Specify the number of participants and outcome events in each analysis (e.g., for model development, hyperparameter tuning, model evaluation) | S23, 30, T1 |
| *Model specification* | 22 | D | Provide details of the full prediction model (e.g., formula, code, object, application programming interface) to allow predictions in new individuals and to enable third-party evaluation and implementation, including any restrictions to access or re-use (e.g., freely available, proprietary)[5] | S12-13, 19* |
| *Model performance* | 23a | D;E | Report model performance estimates with confidence intervals, including for any key subgroups (e.g., sociodemographic). Consider plots to aid presentation. | **S25-26, T2** |
| | 23b | D;E | If examined, report results of any heterogeneity in model performance across clusters. See TRIPOD Cluster for additional details[3]. | NA |
| *Model updating* | 24 | E | Report the results from any model updating, including the updated model and subsequent performance | NA |
| **DISCUSSION** | | | | |
| *Interpretation* | 25 | D;E | Give an overall interpretation of the main results, including issues of fairness in the context of the objectives and previous studies | M15-16* |
| *Limitations* | 26 | D;E | Discuss any limitations of the study (such as a non-representative sample, sample size, overfitting, missing data) and their effects on any biases, statistical uncertainty, and generalizability | M20-21, S14 |
| *Usability of the model in the context of current care* | 27a | D | Describe how poor quality or unavailable input data (e.g., predictor values) should be assessed and handled when implementing the prediction model | S3* |
| | 27b | D | Specify whether users will be required to interact in the handling of the input data or use of the model, and what level of expertise is required of users | M10-11, S15* |
| | 27c | D;E | Discuss any next steps for future research, with a specific view to applicability and generalizability of the model | M20-21 |

From: Collins GS, Moons KGM, Dhiman P, et al. BMJ 2024;385:e078378. doi:10.1136/bmj-2023-078378

M = Manuscript; S = Supplement. T1, T2, T3 = separate Tables 1, 2, 3 (pages 1 - 3, 1 - 2, 1 - 2, respectively).
F2D = Figure 2D. NR = not reported. NA = not applicable. An asterisk indicates partial reporting.

[4] This relates to the analysis code, for example, any data cleaning, feature engineering, model building, evaluation.
[5] This relates to the code to implement the model to get estimates of risk for a new individual.

### Supplementary STARD-AI checklist

| Section and Topic | No. | STARD-AI Item | Reported on page |
|---|---|---|---|
| **Title or abstract** | | | |
| | 1 | Identification as a study reporting AI-centred diagnostic accuracy and reporting at least one measure of accuracy within title or abstract | M3 - 4 |
| **Abstract** | | | |
| | 2 | Structured summary of study design, methods, results and conclusions (for specific guidance, please see STARD for Abstracts) | M3 - 4 |
| **Introduction** | | | |
| | 3 | Scientific and clinical background, including the intended use of the index test, whether it is novel or an established index test, and its integration into an existing or new workflow, if applicable | M6-7, 15-16 |
| | 4 | Study objectives and hypotheses | M7 |
| **Methods** | | | |
| Study design | 5 | Whether data collection was planned before the index test and reference standard were performed (prospective study) or after (retrospective study) | M7-8, 10-11 |
| Ethics | 6 | Formal approval from an ethics committee. If not required, justify why | M8-9 |
| Participants | 7 | Eligibility criteria: listing separate inclusion and exclusion criteria in the order that they are applied at both participant level and data level | M7-8, S2-3 |
| | 8 | On what basis potentially eligible participants were identified (such as symptoms, results from previous tests, inclusion in registry) | M7-8 |
| | 9 | Where and when potentially eligible participants were identified (setting, location, and dates) | M7-8 |
| | 10 | Whether participants formed a consecutive, random, or convenience series | M7-8, 10-11 |
| Dataset | 11 | Source of the data and whether it has been routinely collected, specifically collected for the purpose of the study or acquired from an open-source repository | M7-9, S2-3 |
| | 12 | Who undertook the annotations for the dataset (including experience levels and background) and how (within the same clinical context or in a post-hoc fashion), if applicable | M7-8, S3* |
| | 13 | Devices (manufacturer, model) that were used to capture data; software (with version number) used to engineer the index test, highlighting the intended use | M12, S2-3* |
| | 14 | Data acquisition protocols (e.g. contrast protocol or reconstruction method for medical images) and details of data pre-processing in sufficient detail to allow replication | M8, S2, 12-13 |
| Test methods | 15a | Index test, in sufficient detail to allow replication | M9-10, S3-13, 19* |
| | 15b | How the index test was developed, including any training, validation, testing and external evaluation, detailing sample sizes, when applicable | M8-10, 12-13<br>S3, 7, 12-13, 30 |
| | 15c | Definition of and rationale for test positivity cut-offs or result categories of the index test, distinguishing pre-specified from exploratory | M10, S14* |
| | 15d | The specified end user of the index test and the level of expertise required of users | M10-11, 15-16<br>S27* |
| | 16a | Reference standard, in sufficient detail to allow replication | M7-8, 12-13<br>S3* |

M = Manuscript. S = Supplement. T1, T2, T3 = separate Tables 1, 2, 3 (pages 1-3, 1-2, 1-2, respectively).
F2D = Figure 2D. NR = not reported. NA = not applicable. An asterisk indicates partial reporting.

| | | | |
|---|---|---|---|
| | 16b | Rationale for choosing the reference standard (if alternatives exist) | M7-8* |
| | 16c | Definition of and rationale for test positivity cut-offs or result categories of the reference standard, distinguishing pre-specified from exploratory | M7-8* |
| | 17a | Whether clinical information and reference standard results were available to the performers or readers of the index test | M10-11* |
| | 17b | Whether clinical information and index test results were available to the assessors of the reference standard | NR |
| Analysis | 18 | Methods for estimating or comparing measures of diagnostic accuracy | M10-12, S14-17 |
| | 19 | How indeterminate index test or reference standard results were handled | M10-11, S3* |
| | 20 | How missing data on the index test and reference standard were handled | M11, S3, 18* |
| | 21 | Any analyses of variability in diagnostic accuracy, distinguishing pre-specified from exploratory | M11-12, S14, 18 |
| | 22 | Intended sample size and how it was determined | M7-8, S16-17 |
| | 23 | Details of any performance error analysis, and algorithmic bias and fairness assessments if undertaken | M15, 31* |
| **Results** | | | |
| Participants and dataset | 24 | Flow of participants, using a diagram | M12-13, 30-31* |
| | 25 | Baseline demographic, clinical and technical characteristics of training, validation and test set, if applicable | S27, 30, T1* |
| | 26a | Distribution of severity of disease in those with the target condition | M12-13, T1* |
| | 26b | Distribution of alternative diagnoses in those without the target condition | S23, T1 |
| | 27 | Time interval and any clinical interventions between index test and reference standard | M7-8, 13* |
| | 28 | Whether the datasets represent the distribution of the target condition that one would expect from the intended use population | M7-8, 20-21, S15* |
| | 29 | For external evaluation on an independent dataset, an assessment of how this differs from the training, validation and test sets | M7-9, 20-21, T1 |
| Test results | 30 | Cross tabulation of the index test results (or their distribution) by the results of the reference standard | F2D, S34-35 |
| | 31 | Estimates of diagnostic accuracy and their precision (such as 95% confidence intervals) | S26, 29, T2, T3 |
| | 32 | Any adverse events from performing the index test or the reference standard | NR |
| **Discussion** | | | |
| | 33 | Study limitations, including sources of potential bias, statistical uncertainty, and generalisability | M20-21, S14 |
| | 34 | Implications for practice, including the intended use and clinical role of the index test | M15-16, 21 |
| | 35 | Ethical considerations and adherence to ethical standards associated with the use of the index test and issues of fairness | M8-9, 19-20* |
| **Other information** | | | |
| | 36 | Registration number and name of registry | S18 |
| | 37 | Where the full study protocol can be accessed | S18 |
| | 38 | Sources of funding and other support; role of funders | M22 |
| | 39 | Commercial interests, if applicable | M22 |
| | 40a | Availability of datasets and code; detailing any restrictions on their reuse and repurposing | M22-23, S19* |
| | 40b | Whether outputs are stored, auditable and available for evaluation, if necessary | S13-14, 19* |

M = Manuscript. S = Supplement. T1, T2, T3 = separate Tables 1, 2, 3 (pages 1-3, 1-2, 1-2, respectively).
F2D = Figure 2D. NR = not reported. NA = not applicable. An asterisk indicates partial reporting.
